# Sensitive period-regulating genetic pathways and exposure to adversity shape risk for depression

**DOI:** 10.1101/2021.05.13.21257179

**Authors:** Yiwen Zhu, Min-Jung Wang, Katherine M. Crawford, Juan Carlos Ramírez-Tapia, Alexandre A. Lussier, Kathryn A. Davis, Christiaan de Leeuw, Anne E. Takesian, Major Depressive Disorder Working Group of the Psychiatric Genomics Consortium, Takao K. Hensch, Jordan W. Smoller, Erin C. Dunn

## Abstract

Animal and human studies have documented the existence of developmental windows (or sensitive periods) when experience can have lasting effects in shaping brain structure or function, behavior, and disease risk. Sensitive periods for depression likely arise through a complex interplay of genes and experience, though this possibility has not been explored. We examined the effect of sensitive period-regulating genetic pathways identified in preclinical animal studies, alone and in interaction with socioeconomic disadvantage, a common childhood adversity, on depression risk. Using a translational approach, we: (1) performed gene-set association analyses using summary data from a genome-wide association study of depression (n=807,553) to assess the effects of three gene sets (60 genes) shown in animal studies to regulate sensitive periods; (2) evaluated the developmental expression patterns of these sensitive period-regulating genes using data from BrainSpan (n=31), a transcriptional atlas of postmortem brain samples; and (3) tested gene-by-development interplay by analyzing the combined effect of common variants in sensitive period genes and timing of exposure to socioeconomic disadvantage within a population-based birth cohort (n=6254). The gene set regulating sensitive period *opening* associated with increased depression risk. Notably, six of the 15 genes in this set showed developmentally regulated gene-level expression. A genome-wide polygenic risk score-by-environment analysis showed socioeconomic disadvantage during ages 1-5 years were independently associated with depression risk, but no gene-by-development interactions were found. Genes involved in regulating sensitive periods may be implicated in depression vulnerability and differentially expressed across the life course, though larger studies are needed to identify developmental interplays.

## Introduction

Sensitive periods are stages of heightened plasticity when experience can have particularly strong and enduring effects on brain structure, behavior, and health [1-5]. To date sensitive periods have been most commonly studied with respect to sensory systems and related domains, including vision, hearing, and language learning, in animals [6-8] and humans [9,10]. This research has revealed that sensitive period plasticity occurs through an orchestration of genes and life experiences [11,12]. As shown in **Figure 1** and previously summarized elsewhere [12], robust evidence from *in vivo* experiments with genetically modified mice or rats has shown that several dozen genes regulate the *opening, closing*, and *expression* of sensitive periods in the visual and auditory systems [12-15]. For instance, *opening* genes (e.g., *Bdnf* or *Gad2* [12,16,17]) initiate, accelerate, or delay the onset of sensitive periods by regulating parvalbumin (PV) cell maturation and altering the ratio of excitatory and inhibitory circuit activity. *Closing* genes (e.g., *Acan, Rtn4r*) regulate the formation of perineuronal nets (PNNs), which operate as a “molecular brake” of sensitive period plasticity [18]. *Expression* genes (e.g., *Nr2a* or *Stat1* [19,20]) maintain the duration of sensitive periods by circuit rewiring and consolidation. Beyond the primary sensory cortex, these genetic pathways have been recently implicated in plasticity mechanisms that configure the prefrontal cortical network [21], which regulates cognition and mood [8,18,21]. Thus, alterations in the genetic structure of sensitive period regulation have developmental impacts across brain regions and could give rise to varying levels of psychiatric vulnerability.

**Figure 1.**
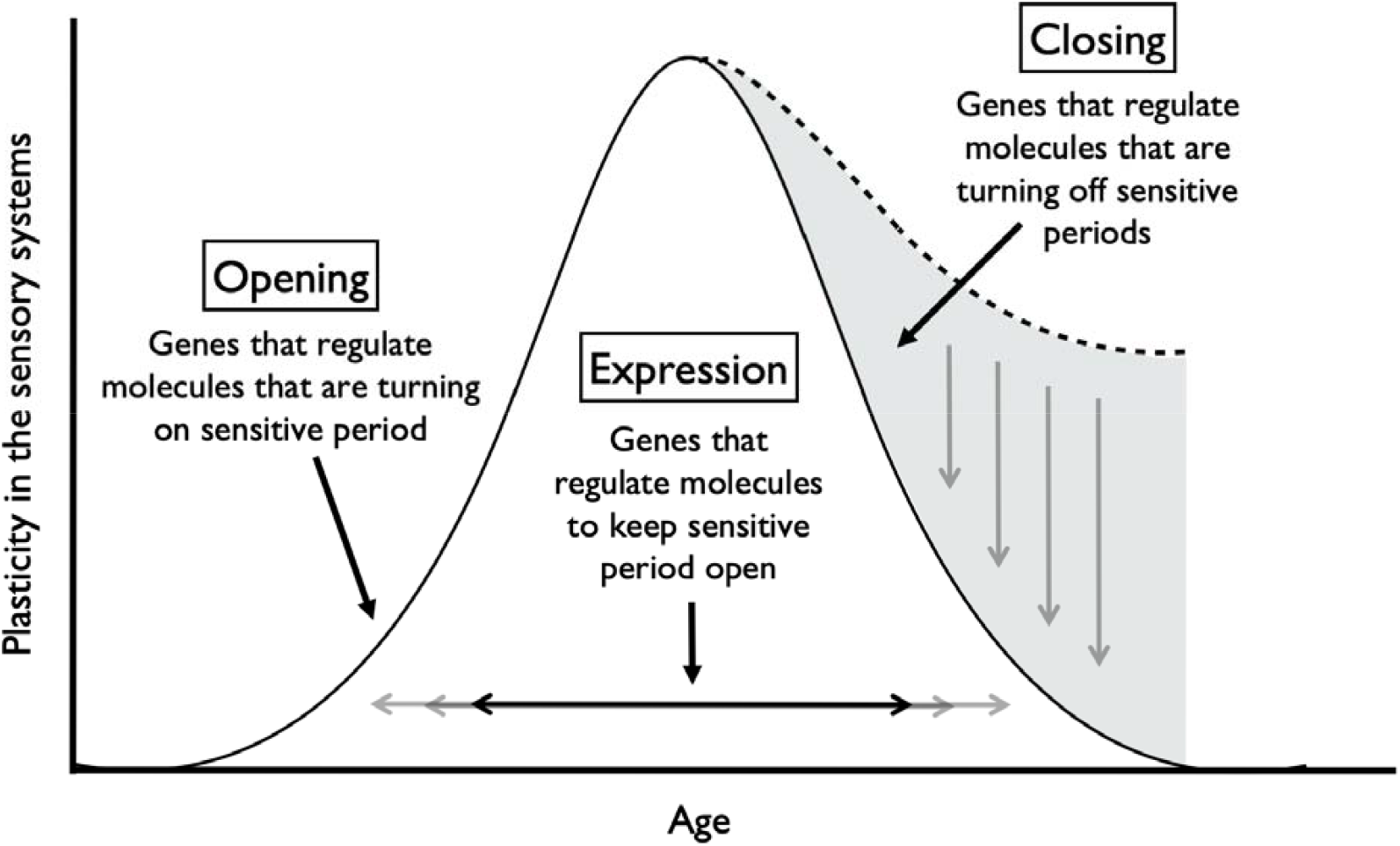
Gene sets regulating three aspects of sensitive period functioning in the sensory systems of mice. *Note*: Gene sets identified by Hensch (2005) and Takesian and Hensch (2013). Opening genes: *BDNF, ARNTL, CLOCK, GABBR1, GABRA1, GABRA2, GABRB3, GAD1, GAD2, SLC6A1, NTRK2, OTX2, NPTX2, HTR3A, CHRNA4* Expression genes: *CREB1, KCNK2, NGF, GRIN2A, DLG4, PVALB, STAT1, TNF* Closing genes: *OTX2, GCLC, GCLM, LYNX1, MAG, MBP, HLA-C, HLA-A, HLA-B, PODN, LILRB3, LILRB1, PTPRS, RTN4, ACAN, BCAN, HAPLN1, HAPLN3, HAPLN4, NCAN, PTS, TNR, VCAN, ADAMTS15, ADAMTS4, ADAMTS8, CSGALNACT1, HAS1, HAS2, HAS3, MME, MMP15, MMP24, MMP3, MMP8, DLG4, PILRB, CAM5, NCAM1*

To that end, accumulating evidence from molecular studies suggest genetic dysregulation of sensitive period plasticity may explain risk for neuropsychiatric disorders. For instance, in mouse models of autism, researchers observed mistimed sensitive period onset due to premature or impoverished PV circuits [24, 25]. In both clinical patients and animal models, reduction of sensitive period triggers (PV+ interneurons) or molecular brakes (PNNs) and sensitive period-related transcriptional or epigenetic aberrations were shown to confer greater risk for schizophrenia [24–26]. Recent findings from large-scale genome-wide association studies (GWAS) also point towards associations between variants located in some of the genes implicated in sensitive period regulation (e.g., *GABBR1* [22-25], *GRIN2A* [26-30], *NCAM1* [23,31], *NCAN* [30]) and neuropsychiatric illnesses, such as schizophrenia, depression, and bipolar disorder.

Environmental perturbations during sensitive periods are also associated with risk for neuropsychiatric disorders [32,33]. Observational epidemiologic studies in humans show that exposure to childhood adversity may have time-dependent effects on brain structure and function [34], as well as social, emotional, and behavioral processes, ranging from fear conditioning to stress reactivity and psychopathology symptoms [4,35,36]. This work has generally found that adversity during the first five years of life is associated with the greatest risk for psychiatric outcomes relative to exposure after age five or no exposure [4]. Adversity is thought to disrupt sensitive period functioning through experience-expectant processes, wherein during restricted periods of development, the brain is primed through genetic instruction to expect a normative set of environmental inputs [37]. Experiences of childhood adversity, including acts of social commission (e.g., physical or sexual abuse) or social and material omission (e.g., neglect, poverty) [38], are therefore understood as violations of expected environmental inputs that can lead to impaired brain plasticity and mental disorders [39].

Depression is a disorder classically viewed as governed by the interplay between genetic variations and time-dependent experiences over the life course [40,41]. While the timing and duration of depression-related sensitive periods are unknown, preclinical and molecular studies have implicated sensitive period biology in etiologic pathways of depression risk. For example, the molecular signature of sensitive period closure, PNNs, has protective effects against oxidative stress [42]. Oxidative stress responses have been connected to the pathophysiology of major depressive disorder [43,44] and appear impacted by childhood adversity [45,46]. Moreover, deficits in *GABA*ergic transmission, which plays a key role in regulating sensitive period timing, can alter susceptibility to early life stress [47]. Therefore, stress exposures during developmental periods of heightened vulnerability may give rise to more severe symptoms of depression, especially among individuals with genetic variations linked to disrupted sensitive period timing [47-49].

However, to our knowledge, no studies to date have investigated the independent and joint roles of gene sets regulating developmental plasticity and exposure to time-varying early life adversity on depression risk. In this study, we pursued the overarching hypothesis that genetic variation governing sensitive period plasticity interacts with adverse life experiences during specific developmental windows to shape risk for depression. We tested this hypothesis using a translational approach, bridging and triangulating sources of evidence (from animal models and human studies) [50], types of data (cross-sectional and longitudinal), and disciplines (genetics, developmental neuroscience, and epidemiology). Adopting a pathway-based approach similar to previous studies on neuropsychiatric disorder etiology [51,52], we focused on three sets of genes encompassing existing molecular evidence for sensitive period biology, which regulate the *opening* (n=15 genes), *closing* (n=39 genes), and *expression* (n=8 genes) of sensitive periods (**Figure 1**). The following three research questions are sequentially investigated (**Figure 2**):

1. Does variation in sensitive period genes identified from animal models predict risk for depression in humans?
2. If so, are the sensitive period genes implicated in depression risk developmentally-regulated? And if so, can we identify variants in these genes that function as developmental expression quantitative trait loci (which we refer to as “d-QTLs)”, shaping developmental timing of expression patterns in the brain?
3. Does variation in sensitive period genes interact with time-dependent effects of adversity to shape levels of depressive symptoms? In other words, is there evidence for developmental gene-environment interplay(dGxE)?

**Figure 2.**
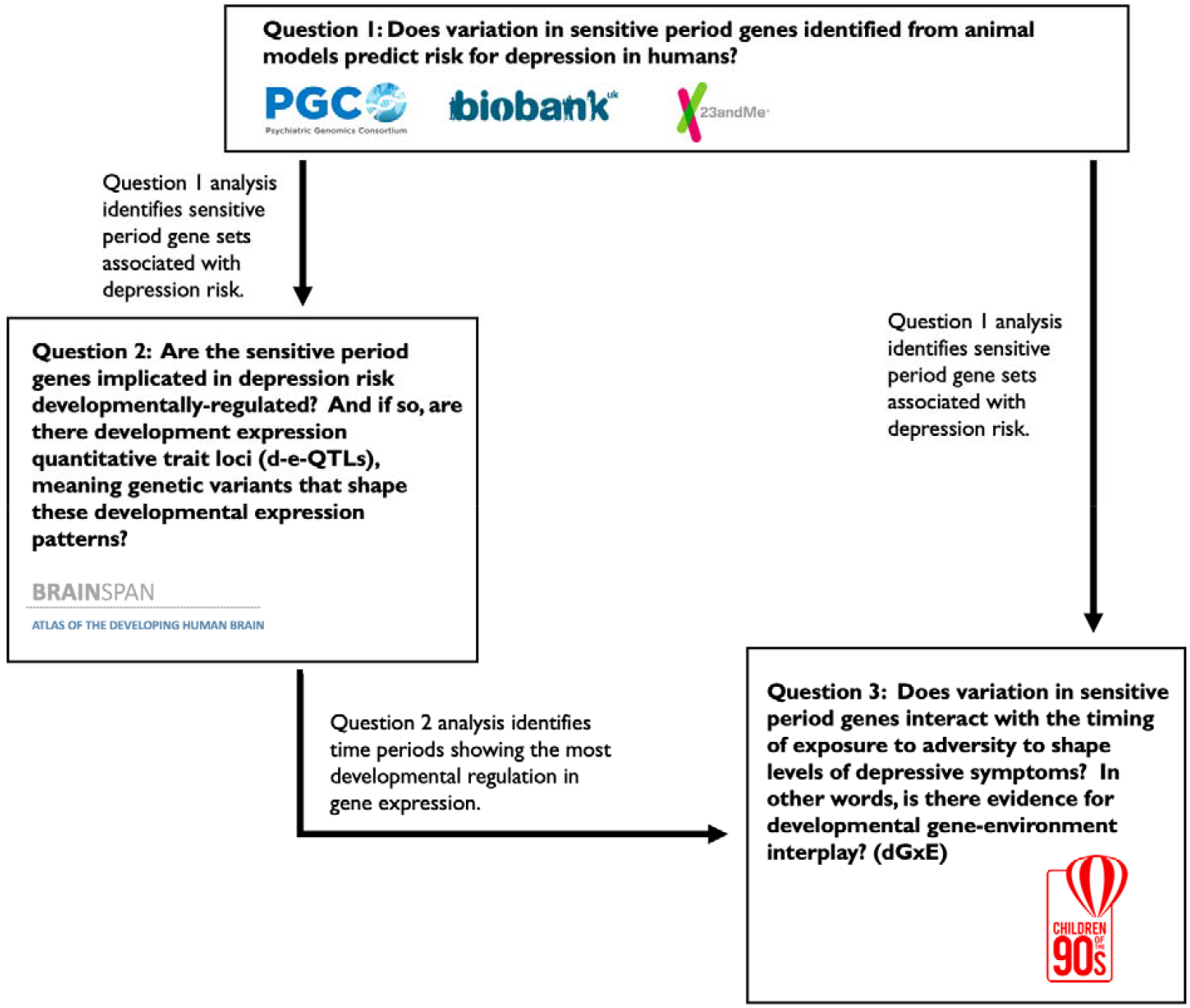
Systematic approach to examining the role of sensitive period-regulating genes, alone and in interaction with exposure to adversity, on risk for depression. *Note:* We approached the above three questions systematically and sequentially, with results from each stage of analysis guiding our approach at the next stage. Specifically, gene sets associated with depression risk in question 1 analysis used to address question 2, where we characterized the developmental trajectories of their gene expression levels and examined whether the trajectories were shaped by certain variants. Similarly, variants annotated to genes in gene sets associated with depression risk were used to compute a gene-set genetic risk score in question 3. Results from question 2 analysis suggested the existence of a biologically-defined sensitive period based on levels of gene expression, which guided how the timing of environmental exposure was parameterized in question 3 analysis.

## Methods and Materials

### Research Question 1: Examining the association between sensitive period gene sets and risk for depression

#### Genetic data

To evaluate the strength of association between genetic pathways involved in regulating sensitive period functioning and risk for depression, we analyzed summary-level data from a meta-analysis of depression, which included 414,055 cases and 892,299 controls from three large-scale depression samples: UK Biobank, the Psychiatric Genomics Consortium (PGC), and 23andMe, Inc. [53]. Across subsamples, depression was defined using minimal (e.g., self-reported symptoms) and deep phenotyping (e.g., structured clinical interviews) approaches. Therefore, the analyses captured genetic architecture of general depression, instead of strict clinical diagnosis of major depressive disorder. Details about the genotyping and quality control procedure are provided by Howard et al. [53].

#### Data Analysis

We pursued a pathway analyses of summary data from the meta-analysis described above. Specifically, we performed competitive gene-set analyses using the Multi-marker Analysis of GenoMic Annotation (MAGMA) software (Version 1.06) [54]. MAGMA uses a nested approach to first summarize SNP-level associations into *gene-level* associations and then *gene-set level* associations, allowing for the detection of aggregated signals at even modest levels. In the current analysis, SNP-level estimates were obtained from the Howard et al. meta-analysis of depression described above [53]. For the *gene-level* analyses, we annotated each of the SNPs reported in the summary data to genes using human genome build 37 (hg19) as the reference. Gene-level p-values were calculated using the sum of -log(p-values) of all tested SNPs within a gene, typically defined as the region between the transcription start and stop sites of each gene [54]. For the *gene-set level* analyses, we grouped the 60 sensitive period genes into their respective sets based on their biological functioning (**Figure 1**). To perform a competitive gene-set analysis, we compared the gene-level results for our three sensitive period gene sets to the gene-level results of the rest of the genome; this test determines whether the average association between genes in the given set and the trait of interest were stronger than that of other genes not in the set. We included a corrected p-value that empirically accounts for multiple testing, using 10,000 random permutations. This gene-set approach, rather than a single genetic variant-level analysis, was chosen to test specific biological pathways and understand their potential for identifying therapeutic targets [54]. Moreover, based on the observation of polygenicity (i.e., many thousands of loci each conferring very small risk), aggregating functionally-consistent signals could substantially improve statistical power [40,55].

Of note, five genes in our analysis were located in the Major Histocompatibility Complex (MHC), a region of the genome involved in human immunity that contains polymorphic loci with long-range linkage-disequilibrium (LD) patterns [56]. Although these features could make interpretation of SNP-level associations more difficult, we did not remove SNPs in the extended MHC region, as LD is explicitly modeled in MAGMA to produce unbiased results (see **Supplemental Materials**).

### Research Question 2: Investigating the developmental regulation of depression-implicated sensitive period gene sets

#### Gene expression data

We investigated the temporal expression patterns of sensitive period genes within the left hemisphere of three brain regions involved in the pathophysiology of depression (amygdala, hippocampus, and medial prefrontal cortex (mPFC)) [57]. Data came from BrainSpan [58,59] (http://www.brainspan.org), a transcriptional atlas of healthy, post-mortem brain donors (ages 5.7 weeks post-conception to 82 years) without large-scale genomic or other abnormalities (see **Supplemental Materials**). In brief, the 31 postnatal donors studied represented both sexes (42% were female) and were predominantly of European ancestry (58%). BrainSpan investigators classified donors by developmental stage at time of death: (1) 0-5 months (n=3); (2) 6-11 months (n=3); (3) 1-5 years (n=2); (4) 6-11 years (n=3); (5) 12-19 years (n=4); (6) 20-39 years (n=9); (7) 40-59 years (n=4); and (8) 60+ (n=3). Prenatal donors and right hemisphere data were not analyzed here, due to issues with data suitability or availability (see **Supplemental Materials**).

Quality-controlled, quantile normalized exon microarray data [60] were downloaded from Gene Expression Omnibus (GEO; GSE25219). For each brain region, expression levels for all probes within an exon were averaged to obtain an expression value for each exon. Probes were annotated to genes using the UCSC Human Genome (hg19) reference sequence. Similar to prior studies [61,62], we used the median of all exons within each gene as the estimate of gene expression. Expression values are presented in log(2) values; thus, each one-unit difference represents a doubling of expression. Although RNA-sequencing data were available in BrainSpan, that dataset contained only one-third of the sample size, which would have been too small for our analysis (see **Supplemental Materials**).

#### Data Analysis

To investigate whether sensitive period gene expression patterns were developmentally regulated, we tested the hypothesis that developmental stage at time of death explained a significant amount of variation in gene expression. Specifically, we performed multiple regression analysis using an omnibus F-test to compare the full model (with developmental stage included as a categorical variable) to a baseline model (only adjusting for two principal components capturing genetic ancestry). The amount of variation in gene expression additionally explained by developmental stage was quantified using the increase in R^2^. To reduce multiple testing burden, we focused only on genes in the gene sets associated with depression risk from analyses of the first research question.

Further, to explore whether certain genetic variants could shape developmental expression patterns of sensitive period genes implicated in depression, we tested the interactions between genotype and developmental stage on expression. Genotype was included as a categorical variable instead of the conventional additive model, to capture the potential nonlinear relationship between genotype and trajectories of gene expression; developmental stage was modeled as an ordinal variable, with both linear and quadratic effects to account for nonlinearity while retaining model parsimony. An F-test was then performed to compare the genotype-by-timing model (with the interaction terms) to the baseline model (without the interaction terms). Of note, we restricted these exploratory analyses to the mPFC, as we saw the most evidence for developmental regulation in this brain region. More details are provided in the **Supplemental Materials**.

### Research Question 3: Investigating interactions between genome-wide and gene set-level genetic liability to depression, timing of exposure to adversity, and depressive symptoms in development (i.e., developmental gene-environment interplay)

#### Dataset and measures

To examine potential developmental gene-environment interplay (dGxE), we analyzed data from 6254 child participants in the Avon Longitudinal Study of Parents and Children (ALSPAC), a UK-based prospective, longitudinal birth-cohort of children followed for more than two decades [63-65]. Ethical approval for the study was obtained from the ALSPAC Ethics and Law Committee and the Local Research Ethics Committees. Consent for biological samples has been collected in accordance with the Human Tissue Act 2004 [66]. More details are available on the ALSPAC website, including a fully searchable data dictionary: http://www.bristol.ac.uk/alspac/researchers/our-data/.

The analytic sample included all participants who were genotyped and had data on the outcome, which was average depressive symptoms across adolescence, spanning ages 10.5 years to 23 years [67,68]. Details about the genotype data collection and quality control procedure are provided in **Supplemental Materials**. Depressive symptoms across adolescence were measured by averaging clinically administered and child self-reports of depressive symptoms using the Short Mood and Feelings Questionnaire (SMFQ) [69] (**Supplemental Materials**). We focused on average symptoms to maximize the analytic sample size and measure general levels of symptoms across development.

We studied socioeconomic disadvantage as our measure of adversity, because it is one of the most commonly-occurring and frequently studied adversity types [70,71], and has been consistently associated with psychopathology symptoms in childhood [72] and adulthood [73,74]. Prior literature [72,75] and our own analyses (see **Supplemental Materials**) provided converging evidence of a time-dependent relationship between exposure to socioeconomic disadvantage and depressive symptoms. In ALSPAC, socioeconomic disadvantage was measured as a time-varying construct based on maternal reports of the extent to which the family had difficulty affording items for the child, rent or mortgage, heating, clothing, or food. Based on prior evidence and preliminary findings, exposure was defined as a three-level variable: no exposure before age 7, exposure during the identified sensitive period (i.e., ages 1-5), and exposure at other time points outside the sensitive period.

#### Data Analysis

Using summary statistics provided by Howard et al. as weights [53], we generated a polygenic risk score (PRS) representing risk for depression conferred by common variants in the gene set(s) associated with depression in the analyses for the first research question, which was comprised of 3617 SNPs before clumping. As a comparison, we additionally generated a genome-wide PRS including all SNPs associated with depression in the summary statistics at the threshold of p<0.05. Prior to computing the PRS, clumping was performed to eliminate SNPs that were in high LD (based on an r^2^ threshold of 0.25). PRS calculations were conducted in PLINK 1.90 [76].

All analyses controlled for the following covariates (measured at childbirth): sex, maternal age, number of previous pregnancies, home ownership, highest level of maternal education, and maternal marital status. We also adjusted for the top four principal components to control for potential population stratification. To reduce potential bias and maximize statistical power, missing exposure and covariate data were multiply imputed using the MICE package [77] in R among participants with complete outcome data. All subsequent multiple regression analyses were performed using 20 imputed datasets and the estimates were combined to account for variations between- and within-imputed datasets.

## Results

### 1) Are genetic pathways involved in regulating sensitive periods associated with depression risk?

Of the three gene sets examined, only the gene set regulating the *opening* of sensitive periods was associated with risk for depression (corrected *p*-value=0.01, **Table 1**). There was no evidence for associations between gene sets involved in the *expression* or *closing* of sensitive periods and depression risk.

**Table 1.** Gene set association analysis for genetic pathways regulating sensitive periods using data from a genome-wide meta-analysis of depression (n=807,553).

| Gene set<br>(sensitive period functioning) | Number of genes | Beta | SE | Unadjusted p-value | Corrected p-value |
| --- | --- | --- | --- | --- | --- |
| opening | 15 | 0.84 | 0.31 | 0.0032 | **0.01** |
| closing | 39 | 0.27 | 0.18 | 0.0656 | 0.1806 |
| expression | 8 | 0.03 | 0.42 | 0.4743 | 0.8493 |

These results came from the most recent genome-wide meta-analysis of depression [53]. Using the Multi-marker Analysis of GenoMic Annotation (MAGMA) software, we first annotated SNPs to genes, ran a gene level analysis, and then ran a competitive gene-set analysis using the gene sets as described, testing the null hypothesis that genes in the tested gene-set were jointly no more associated with the phenotype than genes not in the gene set. The corrected p-value refers to p-values based on 10,000 random permutations, which empirically accounted for multiple testing.

### 2) Are sensitive period genes implicated in depression risk developmentally regulated?

Developmental stage was significantly associated with expression levels in six *opening* genes, explaining up to 54% of the additional variation in gene-level expression beyond genetic ancestry (**Table S1**). We observed preliminary evidence for developmental regulation in the mPFC but not hippocampus or amygdala (**Figure 3)**. Three of the six *opening* genes with evidence of developmental regulation had a nadir of expression between ages 1 and 5: gene expression levels between ages 1 and 5 years were statistically significantly different from other time points at *GABRA1 (*β=-2.33, 95% C.I. [-3.47, -1.19], p=3×10^−4^), *GAD1* (β=-1.88, 95% C.I. [-2.78, -0.98], p=2×10^−4^), and *GAD2* (β=-2.32, 95% C.I. [-3.25, -1.39], p=2×10^−5^).

**Figure 3.**
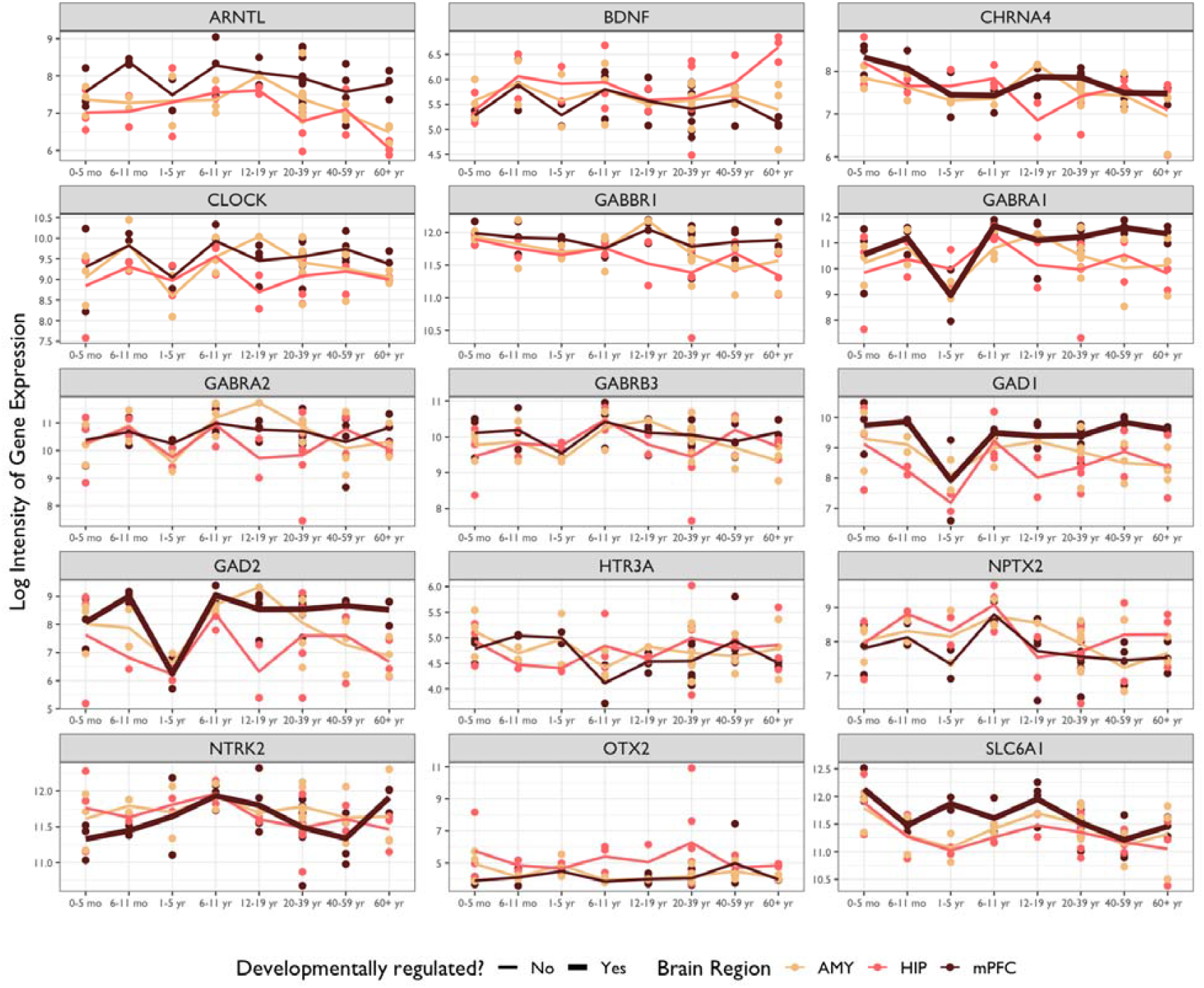
Temporal expression patterns of genes involved in regulating the opening of sensitive periods. *Note:* AMY = Amygdala; HIP = Hippocampus; mPFC = Medial Prefrontal Cortex. Thicker lines indicated a significant association between developmental stage and gene expression (i.e., developmental regulation) based on results from multiple regression analyses. The evidence for developmental regulation was found in the mPFC. In particular, three genes showed decreased expression between ages 1 and 5 (*GABRA1, GAD1*, and *GAD2*).

We further examined whether genotype was associated with developmental expression of these genes. To identify SNPs associated with developmental gene expression (which we term “developmental expression quantitative trait loci”, or d-QTLs), we tested SNP-by-age interactions on expression levels of the *opening* genes. Analyses were limited to the mPFC and examined 144 independent SNPs (**Supplemental Materials**).

We found nominal evidence (*p*<0.05) for SNP-by-age interactions at five loci (**Table S2**), with two SNPs (rs1442060, an intron variant in *GABRA2*; rs7900976, an intron variant in *GAD2*) showing significant association after accounting for the number of SNPs tested within each gene. As shown in **Figure S1**, these results revealed that *GABRA2* was upregulated early in life and downregulated later in life for both major and minor allele homozygotes, whereas the expression level was more stable over time for heterozygotes. *GAD2* was upregulated over time for major allele homozygotes and downregulated for heterozygous individuals. There were only two data points available from homozygous minor individuals, so the trend over time was not discernible. These findings suggest that genetic variants may be associated with different patterns of gene expressions over the life course.

### 3) How does variation in these sensitive period genes interact with the timing of exposure to adversity to shape depressive symptoms?

Average depressive symptoms across adolescence were heritable in our sample (h^2^_SNP_=9.1%, SE=0.05, *p*=0.03). The genome-wide PRS for depression was significantly associated with average depressive symptoms (β=0.40, 95% C.I. [0.31, 0.49], p<1×10^−22^).

The gene set-level PRS representing all SNPs annotated to the *opening* genes did not have a main effect on the outcome (β=-0.01, 95% C.I. [-0.11, 0.08], p=0.78). The environmental exposure, socioeconomic disadvantage, showed a time-dependent association with depressive symptoms. Specifically, the exposure between ages 1 and 5 years was associated with increased symptoms (β=0.79, 95% C.I. [0.5, 1.08], p=1.01×10^−7^). This effect estimate was almost twice as large as the increase associated with exposure at other time points (β=0.43, 95% C.I. [0.07, 0.78], p=0.02). When assessing main genetic and environmental effects additively, we observed similar patterns: both the genome-wide PRS and exposure to socioeconomic disadvantage were associated with depressive symptoms, and no gene set-level PRS effect was detected (**Table 2, Models 4 and 5**). There was no significant evidence supporting dGxE with either the genome-wide PRS or the gene set-level PRS of the *opening* genes (**Table 2, Models 6 and 7**). These findings suggest that there was little signal for gene-by-development interplay, though environmental exposure and genetic risk may jointly shape depression risk additively.

**Table 2.** Effect estimates of main genetic effects, main environmental effect, and GxE interactions using data from ALSPAC (n=6,254).

|  | **Effect estimate** | **Standard error** | **p-value** | **95% CI** |
| --- | --- | --- | --- | --- |
| *Model 1: Main genetic effect, genome-wide PRS* |  |  |  |  |
| Standardized PRS for depression (p<0.05) | 0.40 | 0.05 | <1x10<sup>-22</sup> | (0.31, 0.49) |
| *Model 2: Main genetic effect, opening genes PRS* |  |  |  |  |
| Standardized genetic score of opening genes | -0.01 | 0.05 | 0.78 | (-0.11, 0.08) |
| *Model 3: Main environmental effect* |  |  |  |  |
| Exposure to socioeconomic disadvantage between 1 and 5 years | 0.79 | 0.15 | 1.07x10<sup>-7</sup> | (0.5, 1.08) |
| Exposure at other time points | 0.43 | 0.18 | 0.02 | (0.07, 0.78) |
| *Model 4: Additive genetic and environmental effects, genome-wide PRS* |  |  |  |  |
| Standardized PRS for depression (p<0.05) | 0.39 | 0.05 | 4.44x10<sup>-16</sup> | (0.30, 0.48) |
| Exposure to socioeconomic disadvantage between 1 and 5 years | 0.75 | 0.15 | 4.72x10<sup>-7</sup> | (0.46, 1.04) |
| Exposure at other time points | 0.40 | 0.18 | 0.03 | (0.04, 0.75) |
| *Model 5: Additive genetic and environmental effects, opening genes PRS* |  |  |  |  |
| Standardized genetic score of opening genes | -0.01 | 0.05 | 0.75 | (-0.11, 0.08) |
| Exposure to socioeconomic disadvantage between 1 and 5 years | 0.79 | 0.15 | 9.67x10<sup>-8</sup> | (0.50, 1.08) |
| Exposure at other time points | 0.43 | 0.18 | 0.02 | (0.08, 0.78) |
| *Model 6: Gene and environment interaction, genome-wide PRS* |  |  |  |  |
| Standardized PRS for depression (p<0.05) | 0.37 | 0.05 | 4.20x10<sup>-12</sup> | (0.26, 0.47) |
| Exposure to socioeconomic disadvantage between 1 and 5 years | 0.72 | 0.15 | 1.15x10<sup>-6</sup> | (0.43, 1.01) |
| Exposure at other time points | 0.40 | 0.18 | 0.03 | (0.05, 0.76) |
| PRS for depression x exposure between 1 and 5 years | 0.26 | 0.15 | 0.09 | (-0.03, 0.55) |
| PRS for depression x exposure at other time points | -0.08 | 0.18 | 0.66 | (-0.43, 0.27) |
| *Model 7: Gene and environment interaction, opening genes PRS* |  |  |  |  |
| Standardized PRS of opening genes | -0.04 | 0.05 | 0.40 | (-0.15, 0.06) |
| Exposure to socioeconomic disadvantage between 1 and 5 years | 0.79 | 0.15 | 1.09x10<sup>-7</sup> | (0.50, 1.08) |
| Exposure at other time points | 0.44 | 0.18 | 0.02 | (0.08, 0.79) |
| Opening genes PRS x exposure between 1 and 5 years | 0.14 | 0.15 | 0.35 | (-0.16, 0.44) |
| Opening genes PRS x exposure at other time points | 0.18 | 0.17 | 0.31 | (-0.16, 0.52) |

## Discussion

Three notable findings emerged from our analyses. First, we found that variations in genes governing the onset of sensitive periods were associated with risk for depression at the population level. The mechanism underlying this association remains to be determined. However, we posit that because these genes are known to regulate the initial maturation of inhibitory signaling during early development [6,12], their altered function might therefore delay the onset of sensitive period plasticity. Such mistiming could result in aberrant responses to external stimuli during the process of fear learning and other cognitive functions critical to affective development [78] and thus increase risk for depression and other neuropsychiatric disorders. By contrast, we found no effect on depression of genetic pathways involved in the duration or closure of sensitive periods. The lack of association with *closing* genes is somewhat surprising given previous studies suggesting that PNN maturation plays an instrumental role in the emergence of higher order functioning (e.g., social memory formation) [79], and has protective effects against toxic stress exposures [42]. The lack of evidence from our study could be due to the heterogeneity among genes included in the *closing* set. Among the 39 genes in this set, some relate to the formation of PNNs (e.g., *ACAN, RTN4R*), while others affect myelin and myelin-associated inhibitors that restrict plasticity (e.g., *PIRB*) [12]. Although these 39 genes all regulate the closure of sensitive periods, they appeared to have mixed effects on depression risk: namely, only 11 out of the 39 genes showed a gene-level association with depression risk. We analyzed the entire *closing* set to preserve the biological interpretation of this pathway and avoid cherry-picking, however, testing somewhat heterogeneous genes as one pathway might have diluted the signals. Readers interested in seeing gene-level results are referred to **Table S5**.

Second, we showed that genes involved in sensitive period functioning were developmentally regulated in the mPFC, though not hippocampus or amygdala. Configurations of the prefrontal cortical network during sensitive periods have been implicated in different developmental trajectories and vulnerability to neuropsychiatric disorders, suggesting that plasticity of mPFC may play a prominent role in development [21]. The absence of evidence for developmental regulation in amygdala or hippocampus was unexpected, because the trajectories of GABAergic signaling markers were found to be nonlinear over time in these brain regions within animal models [80]. Our inability to identify time-varying patterns in these regions could be attributed to the limited sample size: although not reaching statistical significance, some genes (e.g., *GABRA1, GAD1*, and *GAD2*) did show similar trends of expression in amygdala and hippocampus. Moreover, the nadir in expression levels of *opening* genes between ages 1 and 5 years suggests that this time period may be developmentally relevant. This decline in plasticity-related transcriptional activities co-occurred with reported periods of decreased synaptogenesis and increased pruning or maturation of the central nervous system, which may signal the start of developing higher-order cognition or more complex behavior [81,82]. However, we were unable to fully disentangle different patterns between individuals across time due to the lack of repeated gene expression measures. Replication studies are needed to further confirm the importance of the developmental window between ages 1 and 5 years. We also provided a *proof of concept* for identifying genetic loci predicting developmental regulation of genes – which we labeled as d-QTLs. Although our results are preliminary and suggestive given the restrictions of sample size and data availability, the d-QTL framework holds potential for studying the heterogeneity of sensitive period timing and responsivity across individuals with different genetic profiles.

Third, in a population-based sample of adolescents, we did not observe any evidence for genetic main effects or gene-by-developmental timing of adversity interplay. However, we did identify a sensitive period effect of exposure to socioeconomic disadvantage, between ages 1 and 5, which is consistent with prior research [72,83]. The lack of genetic associations with sensitive period genes might reflect insufficient power in our study. As environmental perturbations likely increase phenotypic heterogeneities and yield more nuanced effects, detecting gene-by-environment interactions requires larger sample sizes [40,84]. Our sample size was smaller compared to population-based cross-sectional analyses where effect modification of depression PRS by environmental exposures have been observed [85]. Additionally, our data came from a homogeneous sample of European ancestry children in the UK, where most families of participants were relatively socioeconomically advantaged [64]. Thus, we might have been unable to capture large detrimental effects of extreme hardship and their potential interactions with genetic variations.

Several limitations of our study are noted. First, the lack of detailed phenotypic data in BrainSpan made it impossible for us to test whether developmental regulations of genetic pathways were impacted by life experiences, and whether different trajectories may contribute to risk for depression. By studying associations between phenotypic experiences and gene expression levels across time in human, future studies can better link observable environmental risks to underlying molecular changes. Second, all three data sources were comprised of predominantly White populations, restricting the generalizability of our findings. It should remain a priority of neuropsychiatric genetic studies to expand the recruitment and research of non-White samples [86]. Third, as mentioned above, our analyses were limited by sample sizes and characteristics of the samples. To fully address these issues, larger individual studies or meta/mega-analyses including high risk samples and rigorous assessments of phenotypes are warranted in future work.

In conclusion, our study provides a translational model for testing novel neurobiological pathways by triangulating concepts and data from genetics, developmental sciences, and epidemiology. Through the integration of genetic risk, experiences over the life course, and physiological markers across domains of functioning, interdisciplinary translational studies hold great potential to unravel the complexity of depression etiology and identify novel targets for prevention or intervention.

## Data Availability

Summary data of the genome-wide association study referenced in the manuscript are available from the original publication. Please contact the authors for information on how to access data used for other sections of the analyses.

## Funding and Disclosure

Research reported in this publication was supported by the National Institute of Mental Health of the National Institutes of Health under Award Numbers K01MH102403 and R01MH113930 (Dr. Dunn) and K24MH094614 (Dr. Smoller). It was also supported in part by a NARSAD Young Investigator Grant from the Brain & Behavior Research Foundation (Dr. Dunn) and an award from the Jacobs Foundation (Drs. Dunn and Takesian). The Psychiatric Genomics Consortium (PGC) has received major funding from the US National Institute of Mental Health and the US National Institute of Drug Abuse (U01 MH109528 and U01 MH1095320). The content is solely the responsibility of the authors and does not necessarily represent the official views of the National Institutes of Health.

The UK Medical Research Council and Wellcome (Grant ref: 217065/Z/19/Z) and the University of Bristol provide core support for ALSPAC. A comprehensive list of grants funding is available on the ALSPAC website (http://www.bristol.ac.uk/alspac/external/documents/grant-acknowledgements.pdf). GWAS data was generated by Sample Logistics and Genotyping Facilities at Wellcome Sanger Institute and LabCorp (Laboratory Corporation of America) using support from 23andMe, Inc. This publication is the work of the authors and all authors will serve as guarantors for the contents of this paper.

Dr. Smoller is an unpaid member of the Bipolar/Depression Research Community Advisory Panel of 23andMe, Inc., a member of the Leon Levy Foundation Neuroscience Advisory Board and received an honorarium for an internal seminar at Biogen, Inc. He is Principal Investigator of a collaborative study of the genetics of depression and bipolar disorder sponsored by 23andMe, Inc., for which 23andMe, Inc. provides analysis time as in-kind support but no payments. The remaining authors (including the contributing Consortium members) have nothing to disclose.

## Acknowledgments

The authors thank Oliver Hoffman, PhD, formerly of the Harvard Chan Bioinformatics Core, Harvard T.H. Chan School of Public Health, Boston, MA, Meeta Mistry, Virginia Fisher, and Kuang Zheng for their assistance with early data analyses. We are extremely grateful to all the families who took part in this study, the midwives for their help in recruiting them, and the whole ALSPAC team, which includes interviewers, computer and laboratory technicians, clerical workers, research scientists, volunteers, managers, receptionists and nurses. We would also like to thank the research participants and employees of 23andMe, Inc. for making this work possible.

## Author contributions

YZ, JWS, TKH, and ECD conceived and designed the analysis. ECD and contributing authors of the Major Depressive Disorder Working Group of the Psychiatric Genomics Consortium acquired the data. YZ, MJW, KMC, JCRT, and KAD performed the analysis. AAL, CL, AET, TKH, JWS, and ECD provided critical feedback on preliminary results and revised the analysis. YZ, MJW, KMC, and ECD drafted the manuscript. All authors critically reviewed the manuscript, gave their final approval, and agreed to be accountable for all aspects of the work in ensuring that questions related to the accuracy or integrity of any part of the work were appropriately investigated and resolved.

## Notes

### Author Declarations

Ethical approval for the study was obtained from the ALSPAC Ethics and Law Committee and the Local Research Ethics Committees. Consent for biological samples has been collected in accordance with the Human Tissue Act (2004).Informed consent for the use of data collected via questionnaires and clinics was obtained from participants following the recommendations of the ALSPAC Ethics and Law Committee at the time.

## References

1 Bornstein MH. Sensitive periods in development: Structural characteristics and causal interpretations. Psychological Bulletin. 1989;105(2):179–97.

2 Bailey DB, Bruer JT, Symons FJ, Lichtman JW. Critical Thinking about Critical Periods. A Series from the National Center for Early Development and Learning. 2001.

3 Knudsen EI. Sensitive Periods in the Development of the Brain and Behavior. Journal of Cognitive Neuroscience. 2004;16(8):1412–25.

4 Dunn EC, McLaughlin KA, Slopen N, Rosand J, Smoller JW. Developmental timing of child maltreatment and symptoms of depression and suicidal ideation in young adulthood: Results from the National Longitudinal Study on Adolescent Health. Depress Anxiety. 2013;30(10).

5 Uliana DL, Gomes FV, Grace AA. Stress impacts corticoamygdalar connectivity in an age-dependent manner. Neuropsychopharmacology. 2020:1–10.

6 Hensch TK. Critical Period Regulation. Annual Review of Neuroscience. 2004;27(1):549–79.

7 Hooks BM, Chen C. Critical periods in the visual system: changing views for a model of experience-dependent plasticity. Neuron. 2007;56(2):312–26.

8 Yang E-J, Lin EW, Hensch TK. Critical period for acoustic preference in mice. Proc Natl Acad Sci USA. 2012;109 Suppl 2:17213–20.

9 Sharma A, Campbell J, Cardon G. Developmental and cross-modal plasticity in deafness: Evidence from the P1 and N1 event related potentials in cochlear implanted children. Int J Psychophysiol. 2015;95(2):135–44.

10 Hakuta K, Bialystok E, Wiley E. Critical Evidence: A Test of the Critical-Period Hypothesis for Second-Language Acquisition. Psychological Science. 2016.

11 Fox SE, Levitt P, Nelson CA. How the Timing and Quality of Early Experiences Influence the Development of Brain Architecture. Child Dev. 2010;81(1):28–40.

12 Takesian AE, Hensch TK. Balancing Plasticity/Stability Across Brain Development. Progress in Brain Research. 2013. p. 3–34.

13 Hensch TK. Critical period plasticity in local cortical circuits. Nature Reviews Neuroscience. 2005;6(11):877–88.

14 Kaneko M, Stellwagen D, Malenka RC, Stryker MP. Tumor Necrosis Factor-α mediates one component of competitive, experience-dependent plasticity in developing visual cortex. Neuron. 2008;58(5):673–80.

15 Tropea D, Van Wart A, Sur M. Molecular mechanisms of experience-dependent plasticity in visual cortex. Philos Trans R Soc Lond B Biol Sci. 2009;364(1515):341–55.

16 Huang ZJ, Kirkwood A, Pizzorusso T, Porciatti V, Morales B, Bear MF, et al. BDNF Regulates the Maturation of Inhibition and the Critical Period of Plasticity in Mouse Visual Cortex. Cell. 1999;98(6):739–55.

17 Anomal R, Villers-Sidani dE, Merzenich MM, Panizzutti R. Manipulation of BDNF Signaling Modifies the Experience-Dependent Plasticity Induced by Pure Tone Exposure during the Critical Period in the Primary Auditory Cortex. PLOS ONE. 2013;8(5):e64208.

18 Lee HHC, Bernard C, Ye Z, Acampora D, Simeone A, Prochiantz A, et al. Genetic Otx2 mis-localization delays critical period plasticity across brain regions. Molecular Psychiatry. 2017;22(5):680–88.

19 Fagiolini M, Katagiri H, Miyamoto H, Mori H, Grant SGN, Mishina M, et al. Separable features of visual cortical plasticity revealed by N-methyl-d-aspartate receptor 2A signaling. PNAS. 2003;100(5):2854–59.

20 Nagakura I, Wart AV, Petravicz J, Tropea D, Sur M. STAT1 Regulates the Homeostatic Component of Visual Cortical Plasticity via an AMPA Receptor-Mediated Mechanism. J Neurosci. 2014;34(31):10256–63.

21 Guirado R, Perez-Rando M, Ferragud A, Gutierrez-Castellanos N, Umemori J, Carceller H, et al. A Critical Period for Prefrontal Network Configurations Underlying Psychiatric Disorders and Addiction. Front Behav Neurosci. 2020;14.

22 Yu H, Yan H, Li J, Li Z, Zhang X, Ma Y, et al. Common variants on 2p16.1, 6p22.1 and 10q24.32 are associated with schizophrenia in Han Chinese population. Molecular Psychiatry. 2017;22(7):954–60.

23 Nagel M, Jansen PR, Stringer S, Watanabe K, Leeuw dCA, Bryois J, et al. Meta-analysis of genome-wide association studies for neuroticism in 449,484 individuals identifies novel genetic loci and pathways. Nature Genetics. 2018;50(7):920–27.

24 Hall LS, Adams MJ, Arnau-Soler A, Clarke T-K, Howard DM, Zeng Y, et al. Genome-wide meta-analyses of stratified depression in Generation Scotland and UK Biobank. Translational Psychiatry. 2018;8.

25 Grove J, Ripke S, Als TD, Mattheisen M, Walters RK, Won H, et al. Identification of common genetic risk variants for autism spectrum disorder. Nature Genetics. 2019;51(3):431–44.

26 Li Z, Chen J, Yu H, He L, Xu Y, Zhang D, et al. Genome-wide association analysis identifies 30 new susceptibility loci for schizophrenia. Nature Genetics. 2017;49(11):1576–83.

27 Lam M, Hill WD, Trampush JW, Yu J, Knowles E, Davies G, et al. Pleiotropic Meta-Analysis of Cognition, Education, and Schizophrenia Differentiates Roles of Early Neurodevelopmental and Adult Synaptic Pathways. The American Journal of Human Genetics. 2019;105(2):334–50.

28 Periyasamy S, John S, Padmavati R, Rajendren P, Thirunavukkarasu P, Gratten J, et al. Association of Schizophrenia Risk With Disordered Niacin Metabolism in an Indian Genome-wide Association Study. JAMA Psychiatry. 2019;76(10):1026–34.

29 Ikeda M, Takahashi A, Kamatani Y, Momozawa Y, Saito T, Kondo K, et al. Genome-Wide Association Study Detected Novel Susceptibility Genes for Schizophrenia and Shared Trans-Populations/Diseases Genetic Effect. Schizophr Bull. 2019;45(4):824–34.

30 Stahl EA, Breen G, Forstner AJ, McQuillin A, Ripke S, Trubetskoy V, et al. Genome-wide association study identifies 30 loci associated with bipolar disorder. Nature Genetics. 2019;51(5):793–803.

31 Baselmans BML, Jansen R, Ip HF, Dongen vJ, Abdellaoui A, Weijer vdMP, et al. Multivariate genome-wide analyses of the well-being spectrum. Nature Genetics. 2019;51(3):445–51.

32 Johnson FK, Delpech J-C, Thompson GJ, Wei L, Hao J, Herman P, et al. Amygdala hyper-connectivity in a mouse model of unpredictable early life stress. Translational Psychiatry. 2018;8(1):1–14.

33 Peña CJ, Smith M, Ramakrishnan A, Cates HM, Bagot RC, Kronman HG, et al. Early life stress alters transcriptomic patterning across reward circuitry in male and female mice. Nature Communications. 2019;10(1):5098.

34 Andersen SL, Tomada A, Vincow ES, Valente E, Polcari A, Teicher MH. Preliminary Evidence for Sensitive Periods in the Effect of Childhood Sexual Abuse on Regional Brain Development. JNP. 2008;20(3):292–301.

35 Bos K, Zeanah CH, Fox NA, Drury SS, McLaughlin KA, Nelson CA. Psychiatric Outcomes in Young Children with a History of Institutionalization. Harv Rev Psychiatry. 2011;19(1):15–24.

36 Dunn EC, Nishimi K, Powers A, Bradley B. Is developmental timing of trauma exposure associated with depressive and post-traumatic stress disorder symptoms in adulthood? J Psychiatr Res. 2017;84:119–27.

37 Greenough WT, Black JE, Wallace CS. Experience and brain development. Child Dev. 1987;58(3):539–59.

38 Gilbert R, Widom CS, Browne K, Fergusson D, Webb E, Janson S. Burden and consequences of child maltreatment in high-income countries. Lancet. 2009;373(9657):68–81.

39 McLaughlin KA, Sheridan MA, Nelson CA. Neglect as a Violation of Species-Expectant Experience: Neurodevelopmental Consequences. Biological Psychiatry. 2017;82(7):462–71.

40 Dunn EC, Brown RC, Dai Y, Rosand J, Nugent NR, Amstadter AB, et al. Genetic determinants of depression: Recent findings and future directions. Harv Rev Psychiatry. 2015;23(1):1–18.

41 Uher R, Zwicker A. Etiology in psychiatry: embracing the reality of poly-gene-environmental causation of mental illness. World Psychiatry. 2017;16(2):121–29.

42 Do KQ, Cuenod M, Hensch TK. Targeting Oxidative Stress and Aberrant Critical Period Plasticity in the Developmental Trajectory to Schizophrenia. Schizophr Bull. 2015;41(4):835–46.

43 Black CN, Bot M, Scheffer PG, Cuijpers P, Penninx Bwjh. Is depression associated with increased oxidative stress? A systematic review and meta-analysis. Psychoneuroendocrinology. 2015;51:164–75.

44 Liu T, Zhong S, Liao X, Chen J, He T, Lai S, et al. A Meta-Analysis of Oxidative Stress Markers in Depression. PLOS ONE. 2015;10(10):e0138904.

45 Theall KP, Drury SS, Shirtcliff EA. Cumulative Neighborhood Risk of Psychosocial Stress and Allostatic Load in Adolescents. Am J Epidemiol. 2012;176(suppl_7):S164–S74.

46 Drury SS, Theall K, Gleason MM, Smyke AT, De Vivo I, Wong JYY, et al. Telomere length and early severe social deprivation: linking early adversity and cellular aging. Molecular Psychiatry. 2012;17(7):719–27.

47 Luscher B, Shen Q, Sahir N. The GABAergic deficit hypothesis of major depressive disorder. Molecular Psychiatry. 2011;16(4):383–406.

48 Cameron JL, Eagleson KL, Fox NA, Hensch TK, Levitt P. Social Origins of Developmental Risk for Mental and Physical Illness. J Neurosci. 2017;37(45):10783–91.

49 LeMoult J, Humphreys KL, Tracy A, Hoffmeister J-A, Ip E, Gotlib IH. Meta-analysis: Exposure to Early Life Stress and Risk for Depression in Childhood and Adolescence. Journal of the American Academy of Child & Adolescent Psychiatry. 2020;59(7):842–55.

50 Bale TL, Abel T, Akil H, Jr WAC, Moghaddam B, Nestler EJ, et al. The critical importance of basic animal research for neuropsychiatric disorders. Neuropsychopharmacology. 2019;44(8):1349–53.

51 The Schizophrenia Working Group of the Psychiatric Genomics C, Rammos A, Gonzalez LAN, Weinberger DR, Mitchell KJ, Nicodemus KK. The role of polygenic risk score gene-set analysis in the context of the omnigenic model of schizophrenia. Neuropsychopharmacology. 2019;44(9):1562–69.

52 Mota NR, Poelmans G, Klein M, Torrico B, Fernàndez-Castillo N, Cormand B, et al. Cross-disorder genetic analyses implicate dopaminergic signaling as a biological link between Attention-Deficit/Hyperactivity Disorder and obesity measures. Neuropsychopharmacology. 2020;45(7):1188–95.

53 Howard DM, Adams MJ, Clarke TK, Hafferty JD, Gibson J, Shirali M, et al. Genome-wide meta-analysis of depression identifies 102 independent variants and highlights the importance of the prefrontal brain regions. Nat Neurosci. 2019.

54 de Leeuw CA, Mooij JM, Heskes T, Posthuma D. MAGMA: Generalized Gene-Set Analysis of GWAS Data. PLoS computational biology. 2015;11(4).

55 Wang K, Li M, Hakonarson H. Analysing biological pathways in genome-wide association studies. Nature Reviews Genetics. 2010;11(12):843–54.

56 Glanville KP, Coleman JRI, Hanscombe KB, Euesden J, Choi SW, Purves KL, et al. Classical Human Leukocyte Antigen Alleles and C4 Haplotypes Are Not Significantly Associated With Depression. Biological Psychiatry. 2020;87(5):419–30.

57 Heim C, Binder EB. Current research trends in early life stress and depression: Review of human studies on sensitive periods, gene–environment interactions, and epigenetics. Experimental Neurology. 2012;233(1):102–11.

58 Kang HJ, Kawasawa YI, Cheng F, Zhu Y, Xu X, Li M, et al. Spatio-temporal transcriptome of the human brain. Nature. 2011;478(7370):483–89.

59 Gulsuner S, Walsh T, Watts AC, Lee MK, Thornton AM, Casadei S, et al. Spatial and Temporal Mapping of De novo Mutations in Schizophrenia To a Fetal Prefrontal Cortical Network. Cell. 2013;154(3):518–29.

60 Bolstad BM, Irizarry RA, Astrand M, Speed TP. A comparison of normalization methods for high density oligonucleotide array data based on variance and bias. Bioinformatics. 2003;19(2):185–93.

61 Das D, Clark TA, Schweitzer A, Yamamoto M, Marr H, Arribere J, et al. A correlation with exon expression approach to identify cis-regulatory elements for tissue-specific alternative splicing. Nucleic Acids Research. 2007;35(14):4845–57.

62 Laiho A, Elo LL. A Note on an Exon-Based Strategy to Identify Differentially Expressed Genes in RNA-Seq Experiments. PLOS ONE. 2014;9(12):e115964.

63 Boyd A, Golding J, Macleod J, Lawlor DA, Fraser A, Henderson J, et al. Cohort Profile: The ‘Children of the 90s’—the index offspring of the Avon Longitudinal Study of Parents and Children. International Journal of Epidemiology. 2013;42(1):111–27.

64 Fraser A, Macdonald-Wallis C, Tilling K, Boyd A, Golding J, Davey Smith G, et al. Cohort Profile: The Avon Longitudinal Study of Parents and Children: ALSPAC mothers cohort. International Journal of Epidemiology. 2013;42(1):97–110.

65 Northstone K, Lewcock M, Groom A, Boyd A, Macleod J, Timpson N, et al. The Avon Longitudinal Study of Parents and Children (ALSPAC): an update on the enrolled sample of index children in 2019. Wellcome Open Res. 2019;4:51.(2004).

67 Harris PA, Taylor R, Thielke R, Payne J, Gonzalez N, Conde JG. Research electronic data capture (REDCap)—A metadata-driven methodology and workflow process for providing translational research informatics support. Journal of Biomedical Informatics. 2009;42(2):377–81.

68 Harris PA, Taylor R, Minor BL, Elliott V, Fernandez M, O’Neal L, et al. The REDCap consortium: Building an international community of software platform partners. Journal of Biomedical Informatics. 2019;95:103208.

69 Messer SC, Angold A, Costello J, Loeber R, Van Kammen W, Stouthamer-Loeber M. Development of a Short Questionnaire for use in Epidemiological Studies of Depression in Children and Adolescents: Factor Composition and Structure Across Development. International Journal of Methods in Psychiatric Research. 1995;5:251–62.

70 Slopen N, Koenen KC, Kubzansky LD. Childhood adversity and immune and inflammatory biomarkers associated with cardiovascular risk in youth: a systematic review. Brain, behavior, and immunity. 2012;26(2):239–50.

71 Slopen N, Koenen KC, Kubzansky LD. Cumulative Adversity in Childhood and Emergent Risk Factors for Long-Term Health. The Journal of Pediatrics. 2014;164(3):631–38.e2.

72 Dunn EC, Soare TW, Raffeld MR, Busso DS, Crawford KM, Davis KA, et al. What life course theoretical models best explain the relationship between exposure to childhood adversity and psychopathology symptoms: recency, accumulation, or sensitive periods? Psychological medicine. 2018:1–11.

73 McLaughlin KA, Breslau J, Green JG, Lakoma MD, Sampson NA, Zaslavsky AM, et al. Childhood socio-economic status and the onset, persistence, and severity of DSM-IV mental disorders in a US national sample. Social Science & Medicine. 2011;73(7):1088–96.

74 Boe T, Balaj M, Eikemo TA, McNamara CL, Solheim EF. Financial difficulties in childhood and adult depression in Europe. European journal of public health. 2017;27(suppl_1):96–101.

75 Teicher MH, Anderson CM, Polcari A. Childhood maltreatment is associated with reduced volume in the hippocampal subfields CA3, dentate gyrus, and subiculum. PNAS. 2012;109(9):E563–E72.

76 Purcell S, Neale B, Todd-Brown K, Thomas L, Ferreira Manuel AR, Bender D, et al. PLINK: A Tool Set for Whole-Genome Association and Population-Based Linkage Analyses. American Journal of Human Genetics. 2007;81(3):559–75.

77 van Buuren S, Groothuis-Oudshoorn K. mice: Multivariate Imputation by Chained Equations in R. Journal of Statistical Software. 2011;45:urn:issn:1548–7660.

78 Hartley CA, Lee FS. Sensitive Periods in Affective Development: Nonlinear Maturation of Fear Learning. Neuropsychopharmacology. 2015;40(1):50–60.

79 Domínguez S, Rey CC, Therreau L, Fanton A, Massotte D, Verret L, et al. Maturation of PNN and ErbB4 Signaling in Area CA2 during Adolescence Underlies the Emergence of PV Interneuron Plasticity and Social Memory. Cell Reports. 2019;29(5):1099–112.e4.

80 King EC, Pattwell SS, Sun A, Glatt CE, Lee FS. Nonlinear developmental trajectory of fear learning and memory. Annals of the New York Academy of Sciences. 2013;1304(1):62–69.

81 Huttenlocher PR, de Courten C, Garey LJ, Van der Loos H. Synaptogenesis in human visual cortex — evidence for synapse elimination during normal development. Neuroscience Letters. 1982;33(3):247–52.

82 Silbereis John C, Pochareddy S, Zhu Y, Li M, Sestan N. The Cellular and Molecular Landscapes of the Developing Human Central Nervous System. Neuron. 2016;89(2):248–68.

83 Gilman SE, Kawachi I, Fitzmaurice GM, Buka SL. Socioeconomic status in childhood and the lifetime risk of major depression. International Journal of Epidemiology. 2002;31(2):359–67.

84 Marigorta UM, Gibson G. A simulation study of gene-by-environment interactions in GWAS implies ample hidden effects. Front Genet. 2014;5.

85 Coleman JRI, Peyrot WJ, Purves KL, Davis KAS, Rayner C, Choi SW, et al. Genome-wide gene-environment analyses of major depressive disorder and reported lifetime traumatic experiences in UK Biobank. Molecular Psychiatry. 2020;25(7):1430–46.

86 Wojcik GL, Graff M, Nishimura KK, Tao R, Haessler J, Gignoux CR, et al. Genetic analyses of diverse populations improves discovery for complex traits. Nature. 2019;570(7762):514–18.

