## Supplementary Materials for "Sensitive period-regulating genetic pathways and exposure to adversity shape risk for depression"

**Mapping Mouse Gene to Human Orthologs**

We identified the human orthologs of sensitive period genes derived from preclinical animal studies using the National Center for Biotechnology Information (NCBI) Gene and NCBI Homologene databases. Of the 57 murine genes, 55 had equivalent gene symbols for the human/mouse ortholog pair. The mouse gene *Pirb* was mapped to *LILRB3* and *LILRB1*; the MHC-I (major histocompatibility complex class I) corresponded to *HLA-A, HLA-B* and *HLA-C*. The mapping resulted in a list of 60 unique genes for our analysis (**Figure 1**).

**Research Question 1: Examination of the association between sensitive period gene sets and risk for depression**

Data quality control and analysis

The SNP-level associations were directly obtained from summary-level data provided by Howard et al. [1], who performed a meta-analysis of depression using data from the UK Biobank, 23andMe, Inc., and the Psychiatric Genomics Consortium. Their analysis was restricted to European only samples (n=807,553) and overlap between the three subsamples was removed. Sample-specific data quality control procedures are described in detail in the original publication.

To perform the gene-level analysis, both imputed and genotyped SNPs were annotated to genes using the 1000 Genomes reference panel build 37 in MAGMA. As the genes regulating sensitive period functioning were small in size (mean=102.106 kb; median=66.501 kb; min=27.146 kb; max=440.87 kb), we took a conservative approach and imposed no gene window. This conservative approach would ensure that no additional SNPs (outside our genes of interest) would be included. Prior studies have shown that when large windows are used, the type I error rate is inflated, and non-causal gene sets are identified, which could result in the inclusion of distant variants being annotated to the genes or gene sets of interest [2].

Following our gene-level analyses, we performed a competitive gene-set analysis using MAGMA, which compares the gene-level results for our gene sets of interest to the gene-level results of the rest of the genome. The competitive analysis evaluates whether the associations in the gene set are on average greater than those of all other genes in the genome. With a competitive analysis, MAGMA tests the null hypothesis that genes in the tested gene set are jointly more associated with the phenotype compared to genes not in the gene set. By default, MAGMA corrects for the confounding effects of gene size, gene density and mean minor allele count by adding these variables as covariates to the gene-level regression model [3]. Further, MAGMA is able to explicitly account for linkage disequilibrium (LD) within 5 Mb by estimating the covariance structure from an appropriate reference panel, without additional pruning needed. Notably, the range of LD accommodated by MAGMA (5Mb) captured longer-range gene-gene correlations in the extended major histocompatibility complex region (MHC) as well.

**Research Question 2: Investigation of developmental regulation of depression-implicated sensitive period gene sets**

Dataset

We assessed developmental regulation of depression-implicated sensitive period gene sets using data from BrainSpan, a transcriptional atlas of 57 healthy, post-mortem donors [4,5]. Both hemispheres (or the whole brain) were collected for 39 donors; the remaining 18 had either left or right hemisphere only. Causes of death in children included: sudden infant death syndrome (SIDS), accidents (e.g., choking; drowning), respiratory insufficiency, asphyxia, acute myocarditis, homicide, and anaphylaxis.

In the current study, we analyzed left hemisphere expression values from 31 postnatal samples, as right hemisphere data was absent for subjects 6-11 years of age, and gene expression values were only modestly correlated between left and right brain regions (average correlation in gene expression among subjects with data from both hemispheres was estimated to be r=0.51). Prenatal samples were also excluded, as we were primarily interested in the trajectories of gene expression during postnatal development. In our analysis, the number of samples per brain region ranged from 24 (amygdala and hippocampus) to 31 (medial prefrontal cortex).

In BrainSpan, gene expression was assessed using exon microarray (ascertained at single-exon resolution for ~1.4 million exon-level probe sets). Quality-controlled, quantile normalized exon microarray data [6] were downloaded from Gene Expression Omnibus (GEO; <http://www.ncbi.nlm.nih.gov/geo/query/acc.cgi?acc=GSE25219>). For each brain region, expression levels for all probes within an exon were averaged to obtain an expression value for each exon. Probes were assigned to genes according to annotation from the UCSC Human Genome (HG19) reference sequence. Taking an approach similar to prior studies [7,8], we used the median of all exons within each gene as the estimate of gene expression. Expression values are presented in log-2 values; thus, each one-unit difference represents a doubling of expression. Although RNA-sequencing data were available, this data was not analyzed as it was collected on only one-third of the sample and thus created too small of a sample for analysis. Specifically, for the medial prefrontal cortex, which is the brain region that our analysis focused on, the sample size for RNA-sequencing data was n=15, and there was no data available beyond 39 years. Therefore, we based our analysis on microarray data of gene expression to maximize the analytical sample size.

All donors were genotyped using Illumina Omni-2.5 million SNP arrays. Details about the laboratory procedure are described by Kang et al. (2011) [5]. Data processing was performed according to the Infinium HD Assay Super, Automated Protocol for Human Omni 2.5-Quad BeadChip (Illumina). If any chromosomal or large-scale genomic abnormalities were detected, the specimens were removed from the sample. In addition to reported ethnicity by next of kin, genetic ancestry was corroborated and refined by cross-referencing the observed genotype allele frequencies with reference populations available in HapMap III. The samples were assigned to European, African, Asian, Hispanic or mixed ancestry accordingly based on an estimated likelihood for each population. The analytic sample (n=31) consisted of 18 donors of European ancestry, 12 donors of African ancestry, and one donor of Hispanic ancestry.

Data analysis

To determine whether there was differential gene expression in these sensitive period genes across developmental stage, we fitted a linear regression model for each gene. This approach was suitable because the observations were not derived from repeated measures of the same individual and could be assumed to be independently distributed. Developmental stage was encoded as a categorical variable, which is a conservative approach that allowed the relationship between developmental stage and gene expression to be non-linear. An F-test was performed to compare the fitted model including developmental stage to the baseline model, which tested the null hypothesis that there were no differences in gene expression across developmental stages (against the alternative hypothesis that the expression level at least one time point was significantly different). Building from analysis addressing the first research question, we focused only on genes in the gene sets that were associated with depression risk from our first analysis; thus, a total of 15 genes were tested.

To identify variants shaping the gene expression trajectories (i.e., d-QTLs), we focused on the medial prefrontal cortex, as we saw the most evidence for developmental regulation in this brain region. SNPs in the BrainSpan sample were annotated to genes of interest using the 1000 Genomes reference panel build 37. Of note, since no imputation was performed in BrainSpan, substantially fewer SNPs were annotated to each gene. To ensure that the SNPs tested were independent, we performed a clumping procedure similar to LD-based clumping to reduce the number of SNPs that were highly correlated. Because the sample came from mixed populations, it was challenging to perform LD-based clumping with a reference population. To emulate LD-based clumping used by standard software, such as PLINK [9], we took the following steps: first, highly correlated variants were selected by identifying pairs of SNPs with a correlation above 0.6. Then the variant with a higher sample minor allele frequency (MAF) in our sample was retained. After the clumping procedure, we performed multiple regression analysis to test interactions between genotype and developmental stage. The first two principal components representing genetic ancestry were included as covariates.

**Research Question 3: Investigating interactions between genome-wide and gene-set-level genetic liability to depression, timing of exposure to adversity, and depressive symptoms in development (i.e., developmental gene-environment interplay)**

Dataset

Data came from the Avon Longitudinal Study of Parents and Children (ALSPAC), a prospective, longitudinal birth-cohort of children born to mothers who were living in the county of Avon England (120 miles west of London) with estimated delivery dates between April 1991 and December 1992 [10-12]. ALSPAC was designed to increase knowledge of the pathways to health across the lifespan, with an emphasis on genetic and environmental determinants. Approximately 85 percent of eligible pregnant women agreed to participate (N=14,541), and 99% of eligible live births (n=14,062) who were alive at 1 year of age (n=13,988 children) were enrolled. An additional attempt to recruit eligible participants that did not join the study original was made when the oldest children reached approximately 7 years of age. Consequently, data after age 7 included new pregnancies beyond the initial sample (Phase I enrollment) described above. According to the current records maintained at age 24, 456, 262, and 195 additional participants were recruited during Phases II, III and IV respectively, resulting in an additional 913 children being enrolled. The total sample size for analyses using any data collected after the age of seven is therefore 15,454 pregnancies, resulting in 15,589 foetuses. Of these 14,901 were alive at 1 year of age. Response rates to data collection have been good (75% have completed at least one follow-up). Ethical approval for the study was obtained from the ALSPAC Ethics and Law Committee and the Local Research Ethics Committee. More details are available on the ALSPAC website, including a fully searchable data dictionary: <http://www.bristol.ac.uk/alspac/researchers/access/>.

Compared to the ALSPAC sample and the subsample with genetic data available, participants in the analytic sample (n=6254) were more likely to be female. They were born to older mothers who had fewer previous pregnancies, more educated, and more likely to be married (**Table S3**).

Measures

*Depressive symptoms*

Depressive symptoms were assessed using the Short Mood and Feelings Questionnaire (SMFQ) [13,14], a 13-item measure commonly used in population-based studies [15-17]. The SMFQ was assessed through child-completed assessments on seven occasions between childhood and young adulthood; two assessments were collected via clinic appointments (at ages 10.5 and 13.5 years old) and five through questionnaires (at ages 16.5, 18.5, 21, 22, and 23). Although the SMFQ was also assessed on three occasions through maternal reporting, we chose not to incorporate these parent-report assessments, given previous literature documenting varying concordance rates between child self-report versus parent-report of children’s depressive symptoms [18,19], particularly among adolescents [20]. The 13 SMFQ items are rated on a three-point scale (0=not at all, 1=sometimes, or 2= true), capturing the severity of depressive symptoms within the past two weeks. The SMFQ has been shown to correlate highly with questionnaire and interview measures of psychopathology and clinician-rated diagnoses of depression [21-25]. Internal consistency in ALSPAC has been found to be excellent (at least 𝛼=0.80) and reliable across participants of different ages [26].

To maximize the analytic sample size for these analyses, we constructed our analytic sample to comprise children who had complete SMFQ data on at least one out of the seven time points of child-completed assessments (n=6254). For each completed SMFQ assessment time point, we summed across the 13 items to create a total SMFQ score between 0 and 26. For each participant, a final depressive symptom score was then calculated as the average of their SMFQ scores across time. We analyzed this average continuous score to maximize statistical power [27].

*Exposure to socioeconomic disadvantage*

Exposure to adversity was assessed by examining socioeconomic disadvantage measured using maternal reports from mail-in questionnaires. We chose socioeconomic disadvantage as our adversity type of interest because it represents one relatively common exposure [28] that is often used to capture “early life adversity” [29,30], and because previous work has demonstrated the robust associations between poverty and risk for mental disorders [31-33]. Our research group has also identified associations between socioeconomic disadvantage and risk for depression in prior ALSPAC-based studies [34]. To capture socioeconomic disadvantage, mothers indicated using a Likert-type scale (1=not difficult; 2=slightly difficult; 3=fairly difficult; 4=very difficult) the extent to which the family had difficulty affording the following: (a) items for the child; (b) rent or mortgage; (c) heating; (d) clothing; (e) food. Children were coded as exposed if their mothers reported that paying for three or more of these necessities was at least fairly difficult; this cut-point roughly corresponded to the top quartile. Questions about financial stress were assessed at seven time-points: when children were approximately ages 8 months, 1.75 years, 2.75 years, 5 years, 7 years, 11 years, and 18 years. In the current study, we included the measurements between ages 8 months and 7 years to ensure that the exposure preceded the outcome.

*Covariates in Regression Models*

Beyond the technical adjustments described in the main text, we additionally controlled for the following variables, measured at child birth: child sex (0=male, 1=female); maternal age (0=ages 15-19, 1=ages 20-35, 2=age>35); number of previous pregnancies (between 0-3+); homeownership (0=mortgage/own home; 1=rent home; 2=other); highest level of maternal education (1=less than O-level, 2=O-level, 3=A-level, 4=Degree or above); and maternal marital status (0=never married; 1=widowed/divorced/separated; 2=married).

*Genetic Data in ALSPAC*

At age 7, genetic data in the ALSPAC sample were collected from a total of 9912 children at age 7. The genotyping was performed using the Illumina HumanHap550 quad genome-wide SNP genotyping platform by 23andMe, Inc. subcontracting the Wellcome Trust Sanger Institute, Cambridge, UK and the Laboratory Corporation of America, Burlington, NC, US. Raw genotype data failing the following quality control filters were excluded: gender mismatches; minimal or excessive heterozygosity; disproportionate levels of individual missingness (>3%), and insufficient sample replication (IBD < 0.8). After performing multidimensional scaling analysis to estimate population stratification using Hapmap Phase II (release 22) reference populations as comparisons, individuals with non-European ancestry were excluded from the final genetic data sample. Additional variant-level quality controls included removing SNPs with a minor allele frequency of < 1%, a call rate of < 95% or evidence for violations of Hardy-Weinberg equilibrium (p < 5x10-7). Cryptic relatedness was examined using the proportion of identity by descent (IBD > 0.1). Subjects that passed all quality control thresholds were included in the sample during subsequent phasing and genotype imputation. A total number of 9115 subjects and 500,527 SNPs were retained in the final genetic data sample.

Data analysis

*Estimation of SNP heritability*

Using genome-wide genetic data from the ALSPAC, we estimated the genetic variation and covariation using genome-wide complex trait analysis (GCTA) [35]. The GCTA heritability analysis integrates information on all available SNPs and estimates the lower-bound additive genetic variance for the phenotype of interest. We first estimated the genetic relatedness matrix, which provides information on genomic similarity across all genotyped SNPs between pair-wise sets of individuals. An estimate of the phenotypic variance attributable to all genotyped SNPs simultaneously were obtained by fitting genetic-relatedness as a random effect in a mixed linear model using a restricted maximum likelihood (REML) function, under a case-control design [36-38] and controlling for sex and principal components. We reported heritability estimates as the proportion of phenotypic variance explained on the underlying prevalence-transformed liability scale [35]. All analyses were implemented in GCTA and R.

*Multiple imputation*

The analytic sample consisted of children that had at least one child-completed assessment of depressive symptoms between ages 10.5 and 23 years. The multiple imputation of the missing covariates and exposure variables was guided by methodological literature [39-41] as well as prior studies that performed imputation using ALSPAC data [42]. The following variables were allowed to enter the imputation models: all covariate and exposure variables, maternal behavior measures during pregnancy (i.e., drinking, smoking, and drug use), other measures of childhood adversity (i.e., caregiver physical or emotional abuse and sexual or physical by anyone) [34], all behavior and depressive symptom measures between ages 11 and 23 years, and 10 principal components derived from genetic data that account for the structure of population ancestry. For each imputed variable, predictors in the list above correlated with the imputed variable or its missingness status (r<0.1) were entered into the imputation model. As all imputed variables were either binary or polytomous, logistic regressions or multinomial logistic regressions were performed, respectively. For each imputed dataset, 25 iterations were used; convergence and data distribution were examined graphically. All imputation was performed in R using the MICE package [39]. Of note, we explored another software package that was specifically designed for phenotype imputation in genetic studies [43], but found that there was limited variation in the imputed values. Hence MICE was used for imputation in the current study.

*Identifying the time-dependent effect of socioeconomic disadvantage on depression*

To understand the time-dependent effect of exposure to socioeconomic disadvantage on depressive symptoms and determine how best to model the main environmental effect in the gene-by-development analysis, we explored age effects in two ways. First, given the results of gene expression patterns explored in the second research question, which showed a potential downregulation of opening genes between ages 1 and 5 years, we modeled the effect of socioeconomic disadvantage during childhood as a categorical variable with three levels: never exposed; exposed during the sensitive period (between ages 1-5); and exposed at another time point (at 8 months and/or ages 6+). As shown in Model 2 of **Table 2**, analyses of this biological definition revealed that on average, children exposed to socioeconomic disadvantage between ages 1 and 5 years had depressive symptoms scores that were 0.79 points higher than children who were never exposed during childhood (95% C.I. [0.50, 1.08], p=1x10^-7^). Children exposed to adversity outside of the sensitive period also showed elevated depressive symptoms compared to those who were never exposed (β=0.43, 95% C.I. [0.07, 0.78], p=0.02).

Second, we also performed an analysis to identify which of the three biologically defined time periods available explained the largest variation in depressive symptoms. We used a structured life course modeling (SLCMA) developed by Mishra [44] and later extended by Smith [45,46] to simultaneously compare competing theoretical models encoding time-varying exposures, and identify the best-fitting theoretical model given the observed variation in average depressive symptoms. In brief, variables encoding competing life course hypotheses were entered into a least angle regression and the variable that explained the most variation in the outcome was selected. For each selected model, a covariance test was performed, testing the null hypothesis that the variable selected is unassociated with the outcome. The method allowed us to assess if there was a theoretical model of socioeconomic disadvantage exposure that explained a significant amount of variation in depressive symptoms, and if so which model(s) was the most important. Given the observed nadir in gene expression trajectories between ages 1 and 5 years shown in the BrainSpan data (see **Figure 3**), we encoded the exposure to socioeconomic disadvantage before age 7 into three variables: exposure before 1 year, exposure between 1 and 5 years, and exposure between 6 and 7 years. More details about the procedure have been described elsewhere [34].

*Examining gene-by-development interplay*

After selecting the best hypothesis using the SLCMA, we constructed a categorical variable to model the exposure to socioeconomic disadvantage during childhood. The outcome (i.e., the average depressive symptom score) was then regressed on the exposure variable, adjusting for the covariates, to obtain the estimates of the main environmental effects.

To accommodate the complexity of the imputed data, we chose a parsimonious modeling approach for the GxE analyses. After conducting the main effect analyses described above, we then tested for interactions between genetic effects and time-dependent exposures to socioeconomic disadvantage. We also fitted full interaction models including the covariate-by-environment and the covariate-by-gene interaction terms as suggested by Keller [47] to control for potential residual confounding. The full interaction models and the reduced interaction models (i.e., GxE terms only) were compared using the pool.compare function in MICE, which implements an F-test for nested linear regression models in the setting of multiply imputed data analysis [39,48].

Results

Results of a SLCMA model [44-46] provided converging evidence in support of a sensitive period between ages 1 and 5(see **Table S4**). Specifically, exposure between ages 1 and 5 years was selected to be the life course hypothesis best supported by the data (p_covariance test_ = 2.5x10^-7^), and no other exposure hypothesis was subsequently selected. In other words, exposure between ages 1 and 5, a biologically defined sensitive period during which genes involved in regulating the onset of sensitive periods were downregulated, might have a particularly potent impact on depressive symptoms.

In the analyses of gene-by-development interplay considering time-varying effects of exposure to socioeconomic disadvantage, we did not detect any signal for potential interactions. The full interaction models accounting for residual confounding of interactions with baseline covariates yielded the same conclusion. The F-tests revealed that the reduced interaction models had sufficient fit compared to the full interaction models. Thus, we presented the reduced interaction models (omitting interactions with baseline covariates) in **Table 2** as our main analyses.

**Major Depressive Disorder Working Group of the Psychiatric Genomics Consortium**

Naomi R Wray* 1, 2

Stephan Ripke* 3, 4, 5

Manuel Mattheisen* 6, 7, 8

Maciej Trzaskowski 1

Enda M Byrne 1

Abdel Abdellaoui 9

Mark J Adams 10

Esben Agerbo 11, 12, 13

Tracy M Air 14

Till F M Andlauer 15, 16

Silviu-Alin Bacanu 17

Marie Bækvad-Hansen 13, 18

Aartjan T F Beekman 19

Tim B Bigdeli 17, 20

Elisabeth B Binder 15, 21

Julien Bryois 22

Henriette N Buttenschøn 13, 23, 24

Jonas Bybjerg-Grauholm 13, 18

Na Cai 25, 26

Enrique Castelao 27

Jane Hvarregaard Christensen 8, 13, 24

Toni-Kim Clarke 10

Jonathan R I Coleman 28

Lucía Colodro-Conde 29

Baptiste Couvy-Duchesne 2, 30

Nick Craddock 31

Gregory E Crawford 32, 33

Gail Davies 34

Ian J Deary 34

Franziska Degenhardt 35

Eske M Derks 29

Nese Direk 36, 37

Conor V Dolan 9

Erin C Dunn 38, 39, 40

Thalia C Eley 28

Valentina Escott-Price 41

Farnush Farhadi Hassan Kiadeh 42

Hilary K Finucane 43, 44

Jerome C Foo 45

Andreas J Forstner 35, 46, 47, 48

Josef Frank 45

Héléna A Gaspar 28

Michael Gill 49

Fernando S Goes 50

Scott D Gordon 29

Jakob Grove 8, 13, 24, 51

Lynsey S Hall 10, 52

Christine Søholm Hansen 13, 18

Thomas F Hansen 53, 54, 55

Stefan Herms 35, 47

Ian B Hickie 56

Per Hoffmann 35, 47

Georg Homuth 57

Carsten Horn 58

Jouke-Jan Hottenga 9

David M Hougaard 13,18

David M Howard 10, 28

Marcus Ising 59

Rick Jansen 19

Ian Jones 60

Lisa A Jones 61

Eric Jorgenson 62

James A Knowles 63

Isaac S Kohane 64, 65, 66

Julia Kraft 4

Warren W. Kretzschmar 67

Zoltán Kutalik 68, 69

Yihan Li 67

Penelope A Lind 29

Donald J MacIntyre 70, 71

Dean F MacKinnon 50

Robert M Maier 2

Wolfgang Maier 72

Jonathan Marchini 73

Hamdi Mbarek 9

Patrick McGrath 74

Peter McGuffin 28

Sarah E Medland 29

Divya Mehta 2, 75

Christel M Middeldorp 9, 76, 77

Evelin Mihailov 78

Yuri Milaneschi 19

Lili Milani 78

Francis M Mondimore 50

Grant W Montgomery 1

Sara Mostafavi 79, 80

Niamh Mullins 28

Matthias Nauck 81, 82

Bernard Ng 80

Michel G Nivard 9

Dale R Nyholt 83

Paul F O'Reilly 28

Hogni Oskarsson 84

Michael J Owen 60

Jodie N Painter 29

Carsten Bøcker Pedersen 11, 12, 13

Marianne Giørtz Pedersen 11, 12, 13

Roseann E Peterson 17, 85

Erik Pettersson 22

Wouter J Peyrot 19

Giorgio Pistis 27

Danielle Posthuma 86, 87

Jorge A Quiroz 88

Per Qvist 8, 13, 24

John P Rice 89

Brien P. Riley 17

Margarita Rivera 28, 90

Saira Saeed Mirza 36

Robert Schoevers 91

Eva C Schulte 92, 93

Ling Shen 62

Jianxin Shi 94

Stanley I Shyn 95

Engilbert Sigurdsson 96

Grant C B Sinnamon 97

Johannes H Smit 19

Daniel J Smith 98

Hreinn Stefansson 99

Stacy Steinberg 99

Fabian Streit 45

Jana Strohmaier 45

Katherine E Tansey 100

Henning Teismann 101

Alexander Teumer 102

Wesley Thompson 13, 54, 103, 104

Pippa A Thomson 105

Thorgeir E Thorgeirsson 99

Matthew Traylor 106

Jens Treutlein 45

Vassily Trubetskoy 4

Andrés G Uitterlinden 107

Daniel Umbricht 108

Sandra Van der Auwera 109

Albert M van Hemert 110

Alexander Viktorin 22

Peter M Visscher 1, 2

Yunpeng Wang 13, 54, 104

Bradley T. Webb 111

Shantel Marie Weinsheimer 13, 54

Jürgen Wellmann 101

Gonneke Willemsen 9

Stephanie H Witt 45

Yang Wu 1

Hualin S Xi 112

Jian Yang 2, 113

Futao Zhang 1

Volker Arolt 114

Bernhard T Baune 115, 116, 117

Klaus Berger 101

Dorret I Boomsma 9

Sven Cichon 35, 47, 118, 119

Udo Dannlowski 114

EJC de Geus 9, 120

J Raymond DePaulo 50

Enrico Domenici 121

Katharina Domschke 122, 123

Tõnu Esko 5, 78

Hans J Grabe 109

Steven P Hamilton 124

Caroline Hayward 125

Andrew C Heath 89

Kenneth S Kendler 17

Stefan Kloiber 59, 126, 127

Glyn Lewis 128

Qingqin S Li 129

Susanne Lucae 59

Pamela AF Madden 89

Patrik K Magnusson 22

Nicholas G Martin 29

Andrew M McIntosh 10, 34

Andres Metspalu 78, 130

Ole Mors 13, 131

Preben Bo Mortensen 11, 12, 13, 24

Bertram Müller-Myhsok 15, 132, 133

Merete Nordentoft 13, 134

Markus M Nöthen 35

Michael C O'Donovan 60

Sara A Paciga 135

Nancy L Pedersen 22

Brenda WJH Penninx 19

Roy H Perlis 38, 136

David J Porteous 105

James B Potash 137

Martin Preisig 27

Marcella Rietschel 45

Catherine Schaefer 62

Thomas G Schulze 45, 93, 138, 139, 140

Jordan W Smoller 38, 39, 40

Kari Stefansson 99, 141

Henning Tiemeier 36, 142, 143

Rudolf Uher 144

Henry Völzke 102

Myrna M Weissman 74, 145

Thomas Werge 13, 54, 146

Cathryn M Lewis* 28, 147

Douglas F Levinson* 148

Gerome Breen* 28, 149

Anders D Børglum* 8, 13, 24

Patrick F Sullivan* 22, 150, 151,

1, Institute for Molecular Bioscience, The University of Queensland, Brisbane, QLD, AU

2, Queensland Brain Institute, The University of Queensland, Brisbane, QLD, AU

3, Analytic and Translational Genetics Unit, Massachusetts General Hospital, Boston, MA, US

4, Department of Psychiatry and Psychotherapy, Universitätsmedizin Berlin Campus Charité Mitte, Berlin, DE

5, Medical and Population Genetics, Broad Institute, Cambridge, MA, US

6, Department of Psychiatry, Psychosomatics and Psychotherapy, University of Wurzburg, Wurzburg, DE

7, Centre for Psychiatry Research, Department of Clinical Neuroscience, Karolinska Institutet, Stockholm, SE

8, Department of Biomedicine, Aarhus University, Aarhus, DK

9, Dept of Biological Psychology & EMGO+ Institute for Health and Care Research, Vrije Universiteit Amsterdam, Amsterdam, NL

10, Division of Psychiatry, University of Edinburgh, Edinburgh, GB

11, Centre for Integrated Register-based Research, Aarhus University, Aarhus, DK

12, National Centre for Register-Based Research, Aarhus University, Aarhus, DK

13, iPSYCH, The Lundbeck Foundation Initiative for Integrative Psychiatric Research,, DK

14, Discipline of Psychiatry, University of Adelaide, Adelaide, SA, AU

15, Department of Translational Research in Psychiatry, Max Planck Institute of Psychiatry, Munich, DE

16, Department of Neurology, Klinikum rechts der Isar, Technical University of Munich, Munich, DE

17, Department of Psychiatry, Virginia Commonwealth University, Richmond, VA, US

18, Center for Neonatal Screening, Department for Congenital Disorders, Statens Serum Institut, Copenhagen, DK

19, Department of Psychiatry, Vrije Universiteit Medical Center and GGZ inGeest, Amsterdam, NL

20, Virginia Institute for Psychiatric and Behavior Genetics, Richmond, VA, US

21, Department of Psychiatry and Behavioral Sciences, Emory University School of Medicine, Atlanta, GA, US

22, Department of Medical Epidemiology and Biostatistics, Karolinska Institutet, Stockholm, SE

23, Department of Clinical Medicine, Translational Neuropsychiatry Unit, Aarhus University, Aarhus, DK

24, iSEQ, Centre for Integrative Sequencing, Aarhus University, Aarhus, DK

25, Human Genetics, Wellcome Trust Sanger Institute, Cambridge, GB

26, Statistical genomics and systems genetics, European Bioinformatics Institute (EMBL-EBI), Cambridge, GB

27, Department of Psychiatry, Lausanne University Hospital and University of Lausanne, Lausanne, CH

28, Social, Genetic and Developmental Psychiatry Centre, King's College London, London, GB

29, Genetics and Computational Biology, QIMR Berghofer Medical Research Institute, Brisbane, QLD, AU

30, Centre for Advanced Imaging, The University of Queensland, Brisbane, QLD, AU

31, Psychological Medicine, Cardiff University, Cardiff, GB

32, Center for Genomic and Computational Biology, Duke University, Durham, NC, US

33, Department of Pediatrics, Division of Medical Genetics, Duke University, Durham, NC, US

34, Centre for Cognitive Ageing and Cognitive Epidemiology, University of Edinburgh, Edinburgh, GB

35, Institute of Human Genetics, University of Bonn, School of Medicine & University Hospital Bonn, Bonn, DE

36, Epidemiology, Erasmus MC, Rotterdam, Zuid-Holland, NL

37, Psychiatry, Dokuz Eylul University School Of Medicine, Izmir, TR

38, Department of Psychiatry, Massachusetts General Hospital, Boston, MA, US

39, Psychiatric and Neurodevelopmental Genetics Unit (PNGU), Massachusetts General Hospital, Boston, MA, US

40, Stanley Center for Psychiatric Research, Broad Institute, Cambridge, MA, US

41, Neuroscience and Mental Health, Cardiff University, Cardiff, GB

42, Bioinformatics, University of British Columbia, Vancouver, BC, CA

43, Department of Epidemiology, Harvard T.H. Chan School of Public Health, Boston, MA, US

44, Department of Mathematics, Massachusetts Institute of Technology, Cambridge, MA, US

45, Department of Genetic Epidemiology in Psychiatry, Central Institute of Mental Health, Medical Faculty Mannheim, Heidelberg University, Mannheim, Baden-Württemberg, DE

46, Department of Psychiatry (UPK), University of Basel, Basel, CH

47, Department of Biomedicine, University of Basel, Basel, CH

48, Centre for Human Genetics, University of Marburg, Marburg, DE

49, Department of Psychiatry, Trinity College Dublin, Dublin, IE

50, Psychiatry & Behavioral Sciences, Johns Hopkins University, Baltimore, MD, US

51, Bioinformatics Research Centre, Aarhus University, Aarhus, DK

52, Institute of Genetic Medicine, Newcastle University, Newcastle upon Tyne, GB

53, Danish Headache Centre, Department of Neurology, Rigshospitalet, Glostrup, DK

54, Institute of Biological Psychiatry, Mental Health Center Sct. Hans, Mental Health Services Capital Region of Denmark, Copenhagen, DK

55, iPSYCH, The Lundbeck Foundation Initiative for Psychiatric Research, Copenhagen, DK

56, Brain and Mind Centre, University of Sydney, Sydney, NSW, AU

57, Interfaculty Institute for Genetics and Functional Genomics, Department of Functional Genomics, University Medicine and Ernst Moritz Arndt University Greifswald, Greifswald, Mecklenburg-Vorpommern, DE

58, Roche Pharmaceutical Research and Early Development, Pharmaceutical Sciences, Roche Innovation Center Basel, F. Hoffmann-La Roche Ltd, Basel, CH

59, Max Planck Institute of Psychiatry, Munich, DE

60, MRC Centre for Neuropsychiatric Genetics and Genomics, Cardiff University, Cardiff, GB

61, Department of Psychological Medicine, University of Worcester, Worcester, GB

62, Division of Research, Kaiser Permanente Northern California, Oakland, CA, US

63, Psychiatry & The Behavioral Sciences, University of Southern California, Los Angeles, CA, US

64, Department of Biomedical Informatics, Harvard Medical School, Boston, MA, US

65, Department of Medicine, Brigham and Women's Hospital, Boston, MA, US

66, Informatics Program, Boston Children's Hospital, Boston, MA, US

67, Wellcome Trust Centre for Human Genetics, University of Oxford, Oxford, GB

68, Institute of Social and Preventive Medicine (IUMSP), University Hospital of Lausanne, Lausanne, VD, CH

69, Swiss Institute of Bioinformatics, Lausanne, VD, CH

70, Division of Psychiatry, Centre for Clinical Brain Sciences, University of Edinburgh, Edinburgh, GB

71, Mental Health, NHS 24, Glasgow, GB

72, Department of Psychiatry and Psychotherapy, University of Bonn, Bonn, DE

73, Statistics, University of Oxford, Oxford, GB

74, Psychiatry, Columbia University College of Physicians and Surgeons, New York, NY, US

75, School of Psychology and Counseling, Queensland University of Technology, Brisbane, QLD, AU

76, Child and Youth Mental Health Service, Children's Health Queensland Hospital and Health Service, South Brisbane, QLD, AU

77, Child Health Research Centre, University of Queensland, Brisbane, QLD, AU

78, Estonian Genome Center, University of Tartu, Tartu, EE

79, Medical Genetics, University of British Columbia, Vancouver, BC, CA

80, Statistics, University of British Columbia, Vancouver, BC, CA

81, DZHK (German Centre for Cardiovascular Research), Partner Site Greifswald, University Medicine, University Medicine Greifswald, Greifswald, Mecklenburg-Vorpommern, DE

82, Institute of Clinical Chemistry and Laboratory Medicine, University Medicine Greifswald, Greifswald, Mecklenburg-Vorpommern, DE

83, Institute of Health and Biomedical Innovation, Queensland University of Technology, Brisbane, QLD, AU

84, Humus, Reykjavik, IS

85, Virginia Institute for Psychiatric & Behavioral Genetics, Virginia Commonwealth University, Richmond, VA, US

86, Clinical Genetics, Vrije Universiteit Medical Center, Amsterdam, NL

87, Complex Trait Genetics, Vrije Universiteit Amsterdam, Amsterdam, NL

88, Solid Biosciences, Boston, MA, US

89, Department of Psychiatry, Washington University in Saint Louis School of Medicine, Saint Louis, MO, US

90, Department of Biochemistry and Molecular Biology II, Institute of Neurosciences, Biomedical Research Centre (CIBM), University of Granada, Granada, ES

91, Department of Psychiatry, University of Groningen, University Medical Center Groningen, Groningen, NL

92, Department of Psychiatry and Psychotherapy, University Hospital, Ludwig Maximilian University Munich, Munich, DE

93, Institute of Psychiatric Phenomics and Genomics (IPPG), University Hospital, Ludwig Maximilian University Munich, Munich, DE

94, Division of Cancer Epidemiology and Genetics, National Cancer Institute, Bethesda, MD, US

95, Behavioral Health Services, Kaiser Permanente Washington, Seattle, WA, US

96, Faculty of Medicine, Department of Psychiatry, University of Iceland, Reykjavik, IS

97, School of Medicine and Dentistry, James Cook University, Townsville, QLD, AU

98, Institute of Health and Wellbeing, University of Glasgow, Glasgow, GB

99, deCODE Genetics / Amgen, Reykjavik, IS

100, College of Biomedical and Life Sciences, Cardiff University, Cardiff, GB

101, Institute of Epidemiology and Social Medicine, University of Münster, Münster, Nordrhein-Westfalen, DE

102, Institute for Community Medicine, University Medicine Greifswald, Greifswald, Mecklenburg-Vorpommern, DE

103, Department of Psychiatry, University of California, San Diego, San Diego, CA, US

104, KG Jebsen Centre for Psychosis Research, Norway Division of Mental Health and Addiction, Oslo University Hospital, Oslo, NO

105, Medical Genetics Section, CGEM, IGMM, University of Edinburgh, Edinburgh, GB

106, Clinical Neurosciences, University of Cambridge, Cambridge, GB

107, Internal Medicine, Erasmus MC, Rotterdam, Zuid-Holland, NL

108, Roche Pharmaceutical Research and Early Development, Neuroscience, Ophthalmology and Rare Diseases Discovery & Translational Medicine Area, Roche Innovation Center Basel, F. Hoffmann-La Roche Ltd, Basel, CH

109, Department of Psychiatry and Psychotherapy, University Medicine Greifswald, Greifswald, Mecklenburg-Vorpommern, DE

110, Department of Psychiatry, Leiden University Medical Center, Leiden, NL

111, Virginia Institute for Psychiatric & Behavioral Genetics, Virginia Commonwealth University, Richmond, VA, US

112, Computational Sciences Center of Emphasis, Pfizer Global Research and Development, Cambridge, MA, US

113, Institute for Molecular Bioscience; Queensland Brain Institute, The University of Queensland, Brisbane, QLD, AU

114, Department of Psychiatry, University of Münster, Münster, Nordrhein-Westfalen, DE

115, Department of Psychiatry, University of Münster, Münster, DE

116, Department of Psychiatry, Melbourne Medical School, University of Melbourne, Melbourne, AU

117, Florey Institute for Neuroscience and Mental Health, University of Melbourne, Melbourne, AU

118, Institute of Medical Genetics and Pathology, University Hospital Basel, University of Basel, Basel, CH

119, Institute of Neuroscience and Medicine (INM-1), Research Center Juelich, Juelich, DE

120, Amsterdam Public Health Institute, Vrije Universiteit Medical Center, Amsterdam, NL

121, Centre for Integrative Biology, Università degli Studi di Trento, Trento, Trentino-Alto Adige, IT

122, Department of Psychiatry and Psychotherapy, Medical Center - University of Freiburg, Faculty of Medicine, University of Freiburg, Freiburg, DE

123, Center for NeuroModulation, Faculty of Medicine, University of Freiburg, Freiburg, DE

124, Psychiatry, Kaiser Permanente Northern California, San Francisco, CA, US

125, Medical Research Council Human Genetics Unit, Institute of Genetics and Molecular Medicine, University of Edinburgh, Edinburgh, GB

126, Department of Psychiatry, University of Toronto, Toronto, ON, CA

127, Centre for Addiction and Mental Health, Toronto, ON, CA

128, Division of Psychiatry, University College London, London, GB

129, Neuroscience Therapeutic Area, Janssen Research and Development, LLC, Titusville, NJ, US

130, Institute of Molecular and Cell Biology, University of Tartu, Tartu, EE

131, Psychosis Research Unit, Aarhus University Hospital, Risskov, Aarhus, DK

132, Munich Cluster for Systems Neurology (SyNergy), Munich, DE

133, University of Liverpool, Liverpool, GB

134, Mental Health Center Copenhagen, Copenhagen Universtity Hospital, Copenhagen, DK

135, Human Genetics and Computational Biomedicine, Pfizer Global Research and Development, Groton, CT, US

136, Psychiatry, Harvard Medical School, Boston, MA, US

137, Psychiatry, University of Iowa, Iowa City, IA, US

138, Department of Psychiatry and Behavioral Sciences, Johns Hopkins University, Baltimore, MD, US

139, Department of Psychiatry and Psychotherapy, University Medical Center Göttingen, Goettingen, Niedersachsen, DE

140, Human Genetics Branch, NIMH Division of Intramural Research Programs, Bethesda, MD, US

141, Faculty of Medicine, University of Iceland, Reykjavik, IS

142, Child and Adolescent Psychiatry, Erasmus MC, Rotterdam, Zuid-Holland, NL

143, Psychiatry, Erasmus MC, Rotterdam, Zuid-Holland, NL

144, Psychiatry, Dalhousie University, Halifax, NS, CA

145, Division of Epidemiology, New York State Psychiatric Institute, New York, NY, US

146, Department of Clinical Medicine, University of Copenhagen, Copenhagen, DK

147, Department of Medical & Molecular Genetics, King's College London, London, GB

148, Psychiatry & Behavioral Sciences, Stanford University, Stanford, CA, US

149, NIHR Maudsley Biomedical Research Centre, King's College London, London, GB

150, Genetics, University of North Carolina at Chapel Hill, Chapel Hill, NC, US

151, Psychiatry, University of North Carolina at Chapel Hill, Chapel Hill, NC, US

**References**

1	Howard DM, Adams MJ, Clarke TK, Hafferty JD, Gibson J, Shirali M, et al. Genome-wide meta-analysis of depression identifies 102 independent variants and highlights the importance of the prefrontal brain regions. Nat Neurosci. 2019.

2	de Leeuw CA, Neale BM, Heskes T, Posthuma D. The statistical properties of gene-set analysis. Nature Reviews Genetics. 2016;17(6):353-64.

3	de Leeuw CA, Mooij JM, Heskes T, Posthuma D. MAGMA: generalized gene-set analysis of GWAS data. PLoS computational biology. 2015;11(4):e1004219.

4	Gulsuner S, Walsh T, Watts AC, Lee MK, Thornton AM, Casadei S, et al. Spatial and temporal mapping of de novo mutations in schizophrenia to a fetal prefrontal cortical network. Cell. 2013;154(3):518-29.

5	Kang HJ, Kawasawa YI, Cheng F, Zhu Y, Zu X, Li M, et al. Spatio-temporal transcriptome of the human brain. Nature. 2011;478:483-89.

6	Bolstad BM, Irizarry RA, Astrand M, Speed TP. A comparison of normalization methods for high density oligonucleotide array data based on variance and bias. Bioinformatics. 2003;19(2):185-93.

7	Das D, Clark TA, Schweitzer A, Yamamoto M, Marr H, Arribere J, et al. A correlation with exon expression approach to identify cis-regulatory elements for tissue-specific alternative splicing. Nucleic Acids Research. 2007;35(14):4845-57.

8	Laiho A, Elo LL. A Note on an Exon-Based Strategy to Identify Differentially Expressed Genes in RNA-Seq Experiments. PLOS ONE. 2014;9(12):e115964.

9	Purcell S, Neale B, Todd-Brown K, Thomas L, Ferreira MAR, Bender D, et al. PLINK: a toolset for whole-genome association and population-based linkage analysis. American Journal of Human Genetics. 2007;81(3):559-75.

10	Boyd A, Golding J, Macleod J, Lawlor DA, Fraser A, Henderson J, et al. Cohort profile: the ‘children of the 90s’—the index offspring of the Avon Longitudinal Study of Parents and Children. International journal of epidemiology. 2012:dys064.

11	Fraser A, Macdonald-Wallis C, Tilling K, Boyd A, Golding J, Davey Smith G, et al. Cohort profile: The Avon Longitudinal Study of Parents and Children: ALSPAC mothers cohort. International Journal of Epidemiology. 2012.

12	Paternoster L, Standl M, Chen C-M, Ramasamy A, Bønnelykke K, Duijts L, et al. Meta-analysis of genome-wide association studies identifies three new risk loci for atopic dermatitis. Nature genetics. 2012;44(2):187-92.

13	Angold A, Costello EJ, Pickles A, Messer SC, Winder F, Silva D. The development of a short questionnaire for use in epidemiological studies of depression in chidlren and adolescents. International Journal of Methods in Psychiatric Research. 1995;5:237-49.

14	Messer SC, Angold A, Costello J, Loeber R, Van Kammen W, Stouthamer-Loeber M. Development of a Short Questionnaire for use in Epidemiological Studies of Depression in Children and Adolescents: Factor Composition and Structure Across Development. International Journal of Methods in Psychiatric Research. 1995;5:251-62.

15	Lundervold AJ, Hinshaw SP, Sorensen L, Posserud MB. Co-occurring symptoms of attention deficit hyperactivity disorder (ADHD) in a population-based sample of adolescents screened for depression. BMC Psychiatry. 2016;16:46.

16	Patton GC, Olsson C, Bond L, Toumbourou JW, Carlin JB, Hemphill SA, et al. Predicting female depression across puberty: a two-nation longitudinal study. J Am Acad Child Adolesc Psychiatry. 2008;47(12):1424-32.

17	Zavos HM, Rijsdijk FV, Gregory AM, Eley TC. Genetic influences on the cognitive biases associated with anxiety and depression symptoms in adolescents. J Affect Disord. 2010;124(1-2):45-53.

18	Achenbach TM, McConaughy SH, Howell CT. Child/adolescent behavioral and emotional problems: Implications of cross-informant correlations for situational specificity. Psychological Bulletin. 1987;101(2):213-32.

19	Grills AE, Ollendick TH. Issues in parent-child agreement: the case of structured diagnostic interviews. Clinical child and family psychology review. 2002;5(1):57-83.

20	Bidaut-Russell M, Reich W, Cottler LB, Robins LN, Compton WM, Mattison RE. The Diagnostic Interview Schedule for Children (PC-DISC v.3.0): Parents and Adolescents Suggest Reasons for Expecting Discrepant Answers. Journal of Abnormal Child Psychology. 1995;23(5):641-59.

21	McKenzie DP, Toumbourou JW, Forbes AB, Mackinnon AJ, McMorris BJ, Catalano RF, et al. Predicting future depression in adolescents using the Short Mood and Feelings Questionnaire: a two-nation study. J Affect Disord. 2011;134(1-3):151-9.

22	Rhew IC, Simpson K, Tracy M, Lymp J, McCauley E, Tsuang D, et al. Criterion validity of the Short Mood and Feelings Questionnaire and one- and two-item depression screens in young adolescents. Child Adolesc Psychiatry Ment Health. 2010;4(1):8.

23	Sharp C, Goodyer IM, Croudace TJ. The Short Mood and Feelings Questionnaire (SMFQ): a unidimensional item response theory and categorical data factor analysis of self-report ratings from a community sample of 7-through 11-year-old children. J Abnorm Child Psychol. 2006;34(3):379-91.

24	Turner N, Joinson C, Peters TJ, Wiles N, Lewis G. Validity of the Short Mood and Feelings Questionnaire in late adolescence. Psychological assessment. 2014;26(3):752-62.

25	Kovacs M. (ed Medicine UoPSo) (University of Pittsburgh School of Medicine, 1983).

26	Stringaris A, Lewis G, Maughan B. Developmental pathways from childhood conduct problems to early adult depression: findings from the ALSPAC cohort. Br J Psychiatry. 2014;205(1):17-23.

27	Otowa T, Hek K, Lee M, Byrne EM, Mirza SS, Nivard MG, et al. Meta-analysis of genome-wide association studies of anxiety disorders. Mol Psychiatry. 2016;21(10):1391-99.

28	Smeeding T, Thévenot C. Addressing Child Poverty: How Does the United States Compare With Other Nations? Academic Pediatrics. 2016;16(3):S67-S75.

29	Slopen N, Koenen KC, Kubzansky LD. Childhood adversity and immune and inflammatory biomarkers associated with cardiovascular risk in youth: a systematic review. Brain, behavior, and immunity. 2012;26(2):239-50.

30	Slopen N, Koenen KC, Kubzansky LD. Cumulative adversity in childhood and emergent risk factors for long-term health. Journal of Pediatrics. 2014;164(3):631-38.

31	Evans GW. The environment of childhood poverty. American Psychologist. 2004;59(2):77-92.

32	Boe T, Balaj M, Eikemo TA, McNamara CL, Solheim EF. Financial difficulties in childhood and adult depression in Europe. European journal of public health. 2017;27(suppl_1):96-101.

33	McLaughlin KA, Breslau J, Green JG, Lakoma MD, Sampson NA, Zaslavsky AM, et al. Childhood socio-economic status and the onset, persistence, and severity of DSM-IV mental disorders in a US national sample. Social Science & Medicine. 2011;73(7):1088-96.

34	Dunn EC, Soare TW, Raffeld MR, Busso DS, Crawford KM, Davis KA, et al. What life course theoretical models best explain the relationship between exposure to childhood adversity and psychopathology symptoms: recency, accumulation, or sensitive periods? Psychological medicine. 2018:1-11.

35	Yang J, Lee SH, Goddard ME, Visscher PM. GCTA: A tool for genome-wide complex trait analysis. American Journal of Human Genetics. 2011;88:76-82.

36	Lee Sang h, Wray Naomi r, Goddard Michael e, Visscher Peter m. Estimating Missing Heritability for Disease from Genome-wide Association Studies. The American Journal of Human Genetics. 2011;88(3):294-305.

37	Lee SH, Yang J, Goddard ME, Visscher PM, Wray NR. Estimation of pleiotropy between complex diseases using single-nucleotide polymorphism-derived genomic relationships and restricted maximum likelihood. Bioinformatics. 2012;28(19):2540-2.

38	Yang J, Benyamin B, McEvoy BP, Gordon S, Henders AK, Nyholt DR, et al. Common SNPs explain a large proportion of the heritability for human height. Nat Genet. 2010;42(7):565-9.

39	van Buuren S, Groothuis-Oudshoorn K. mice: Multivariate Imputation by Chained Equations in R. Journal of Statistical Software. 2011;45:urn:issn:1548-7660.

40	Azur MJ, Stuart EA, Frangakis C, Leaf PJ. Multiple imputation by chained equations: what is it and how does it work? International Journal of Methods in Psychiatric Research. 2011;20(1):40-9.

41	White IR, Royston P, Wood AM. Multiple imputation using chained equations: issues and guidance for practice. Statistics in Medicine. 2011;30(4):377-99.

42	Dunn EC, Soare TW, Raffeld MR, Busso DS, Crawford KM, Davis KA, et al. What life course theoretical models best explain the relationship between exposure to childhood adversity and psychopathology symptoms: recency, accumulation, or sensitive periods? Psychological Medicine. Epub ahead of print:1.

43	Dahl A, Iotchkova V, Baud A, Johansson A, Gyllensten U, Soranzo N, et al. A multiple-phenotype imputation method for genetic studies. Nat Genet. 2016;48(4):466-72.

44	Mishra G, Nitsch D, Black S, De Stavola B, Kuh D, Hardy R. A structured approach to modelling the effects of binary exposure variables over the life course. International Journal of Epidemiology. 2009;38(2):528-37.

45	Smith AD, Heron J, Mishra G, Gilthorpe MS, Ben-Shlomo Y, Tilling K. Model Selection of the Effect of Binary Exposures over the Life Course. Epidemiology. 2015;26(5):719-26.

46	Smith AD, Hardy R, Heron J, Joinson CJ, Lawlor DA, Macdonald-Wallis C, et al. A structured approach to hypotheses involving continuous exposures over the life course. Int J Epidemiol. 2016;45(4):1271-79.

47	Keller MC. Gene × Environment Interaction Studies Have Not Properly Controlled for Potential Confounders: The Problem and the (Simple) Solution. Biological Psychiatry. 2014;75(1):18-24.

48	Meng X-L, Rubin DB. Performing Likelihood Ratio Tests with Multiply-Imputed Data Sets. Biometrika. 1992;79(1):103-11.

| Table S1 Differential gene expression across developmental stage in BrainSpan among genes involved in regulating the opening of sensitive periods. | | | | | | |
| --- | --- | --- | --- | --- | --- | --- |
| Gene | Hippocampus | | Amygdala | | Medial Prefrontal Cortex | |
|  | p-value | Increased R^2^ | p-value | Increased R^2^ | p-value | Increased R^2^ |
| GABBR1 | 0.4419 | 0.342 | 0.4676 | 0.333 | 0.5879 | 0.211 |
| GABRA2 | 0.539 | 0.308 | 0.0917 | 0.507 | 0.6293 | 0.198 |
| GABRB3 | 0.5086 | 0.316 | 0.2561 | 0.41 | 0.667 | 0.181 |
| **GAD1** | 0.3679 | 0.331 | 0.6299 | 0.264 | **0.0128** | **0.52** |
| **GAD2** | 0.6236 | 0.26 | 0.0693 | 0.512 | **0.0024** | **0.535** |
| **SLC6A1** | 0.2682 | 0.393 | 0.4975 | 0.311 | **0.0175** | **0.485** |
| NPTX2 | 0.1866 | 0.387 | 0.1932 | 0.446 | 0.1698 | 0.353 |
| CLOCK | 0.8176 | 0.186 | 0.4596 | 0.32 | 0.6194 | 0.161 |
| **GABRA1** | 0.8543 | 0.179 | 0.5329 | 0.292 | **0.0154** | **0.461** |
| **NTRK2** | 0.6115 | 0.21 | 0.9855 | 0.079 | **0.0368** | **0.464** |
| OTX2 | 0.8388 | 0.19 | 0.2726 | 0.353 | 0.0725 | 0.409 |
| **CHRNA4** | 0.1629 | 0.462 | 0.1937 | 0.419 | **0.0204** | **0.504** |
| P-values were obtained from F-tests comparing models with j categorical developmental stage adjusting for covariates to the covariate-only model. The approach tested the hypothesis: b1=b2=…=bj=0. Compared to ordinal coding, the categorical coding of developmental stage was chosen to capture any presence of nonlinear differential gene expression across ages.  Bolded values indicate genes differentially expressed based on the significance threshold p<0.05. Increased R^2^ values were obtained by subtracting the variance explained by covariate-only model from the variance explained by the age + covariates model, to reflect the additional variation in gene expression explained by developmental stage. | | | | | | |

| Table S2. F-statistics and p-values testing the interactions between developmental stage and SNP-level genetic variation (d-QTLs) in BrainSpan. | | | | |
| --- | --- | --- | --- | --- |
| **Gene** | **Number of SNPs tested** | **SNP** | **F-statistic** | **p-value** |
| GABRA1 | 3 | rs4367330 | 3.84 | 0.0179 |
| **GABRA2** | **9** | **rs1442060** | **11.14** | **0.0001** |
|  |  | rs16859306 | 4.14 | 0.0313 |
| GABRB3 | 39 | rs12324292 | 3.79 | 0.0188 |
|  |  | rs6576603 | 4.01 | 0.0336 |
|  |  | rs878960 | 3.41 | 0.0279 |
| **GAD2** | **2** | **rs7900976** | **9.48** | **0.0004** |
| NTRK2 | 35 | rs1187350 | 3.43 | 0.0273 |
|  |  | rs2277192 | 4.88 | 0.0182 |
|  |  | rs2808707 | 3.89 | 0.0171 |
|  |  | rs7046974 | 4.06 | 0.0308 |
| Two SNPs (rs1442060 and rs7900976) showed significant interactions with developmental stage after correcting for multiple testing at the gene level (α=0.05/number of SNPs tested). In particular, the interaction effect at rs1442060 was still significant after a more stringent Bonferroni correction accounting for the total number of SNPs tested (α=0.05/144=0.0003). | | | | |

| Table S3. Sociodemographic characteristics of the analytic sample (N=6254) compared to the entire ALSPAC sample and genetic subsample. | | | | | |
| --- | --- | --- | --- | --- | --- |
|  | **ALSPAC  (N=15445)** | **Genetic Subsample  (N= 8082)** | **Analytic Sample  (N=6254)** | **ALSPAC vs. Analytic** | **Genetic Subsample vs. Analytic** |
|  | n (%) | n (%) | n (%) | χ2 test  p-value | χ2 test  p-value |
| Sex |  |  |  | <0.001 | <0.001 |
| Males | 7542 (51.3) | 4138 (51.2) | 3026 (48.4) |  |  |
| Females | 7152 (48.7) | 3944 (48.8) | 3228 (51.6) |  |  |
| Age of Mother at Child's Birth |  |  |  | <0.001 | <0.001 |
| Ages 15-19 | 650 (4.6) | 211 (2.7) | 103 (1.7) |  |  |
| Ages 20-35 | 12363 (88.4) | 6912 (89.4) | 5356 (89.8) |  |  |
| Age 36+ | 968 (6.9) | 605 (7.8) | 507 (8.5) |  |  |
| Number of previous pregnancies |  |  |  | <0.001 | <0.001 |
| 0 | 5800 (44.7) | 3316 (44.8) | 2657 (45.8) |  |  |
| 1 | 4550 (35.0) | 2688 (36.3) | 2106 (36.3) |  |  |
| 2 | 1860 (14.3) | 1018 (13.8) | 769 (13.3) |  |  |
| 3+ | 772 (5.9) | 378 (5.1) | 266 (4.6) |  |  |
| Maternal Education |  |  |  | <0.001 | <0.001 |
| less than O-level | 3735 (30.0) | 1821 (25.0) | 1231 (21.3) |  |  |
| O-level | 4303 (34.6) | 2533 (34.8) | 2021 (35.0) |  |  |
| A-level | 2795 (22.5) | 1807 (24.8) | 1542 (26.7) |  |  |
| Degree or Above | 1603 (12.9) | 1114 (15.3) | 978 (16.9) |  |  |
| Maternal Marital Status |  |  |  | <0.001 | <0.001 |
| Never Married | 2522 (19.2) | 1141 (15.3) | 757 (12.9) |  |  |
| Widowed/Divorced/Separated | 787 (6.0) | 410 (5.5) | 283 (4.8) |  |  |
| Married | 9838 (74.8) | 5920 (79.2) | 4815 (82.2) |  |  |

| Table S4. Results of the structured life course modeling approach (SLCMA) examining the association between exposure to socioeconomic disadvantage and depressive symptoms in the ALSPAC analytic sample (n=6254). | | | | | |
| --- | --- | --- | --- | --- | --- |
| Life course hypothesis selected | Increase in R^2^ | Covariance test p-value | Beta (Effect estimate) | SE | 95% CI |
| Exposure between 1 and 5 years | 0.34% | 2.5x10^-7^ | 0.75 | 0.15 | (0.46, 1.04) |
| Exposure before 1 year | 0.34% | 0.93 | - | - | - |
| Exposure between 6 and 7 years | 0.45% | 0.26 | - | - | - |

| Table S5. Gene -level associations between genes regulating sensitive periods and risk for depression, using data from a genome-wide meta-analysis of depression (n=807,553). | | | | | | | | |
| --- | --- | --- | --- | --- | --- | --- | --- | --- |
| **Gene set** | **Gene Name** | **Entrez ID** | **Chromosome** | **Start position** | **Stop position** | **Number of SNPs** | ***Z*-statistic** | ***P*-value** |
| opening | GAD1 | 2571 | 2 | 171673200 | 171717661 | 107 | -0.42 | 6.6E-01 |
|  | SLC6A1 | 6529 | 3 | 11034420 | 11080935 | 157 | -0.25 | 6.0E-01 |
|  | GABRA2 | 2555 | 4 | 46251575 | 46392056 | 343 | -0.07 | 5.3E-01 |
|  | **CLOCK** | **9575** | **4** | **56298660** | **56412997** | **417** | **3.95** | **3.9E-05** |
|  | **GABRA1** | **2554** | **5** | **161274197** | **161326965** | **148** | **4.35** | **6.7E-06** |
|  | GABBR1 | 2550 | 6 | 29570005 | 29600962 | 8 | -1.53 | 9.4E-01 |
|  | NPTX2 | 4885 | 7 | 98246597 | 98259181 | 25 | -0.88 | 8.1E-01 |
|  | **NTRK2** | **4915** | **9** | **87283466** | **87638505** | **988** | **4.12** | **1.9E-05** |
|  | GAD2 | 2572 | 10 | 26505236 | 26593491 | 215 | 1.45 | 7.4E-02 |
|  | **ARNTL** | **406** | **11** | **13299325** | **13408813** | **269** | **2.16** | **1.5E-02** |
|  | **BDNF** | **627** | **11** | **27676440** | **27743605** | **123** | **1.72** | **4.2E-02** |
|  | HTR3A | 3359 | 11 | 113845797 | 113861035 | 52 | 1.17 | 1.2E-01 |
|  | **OTX2** | **5015** | **14** | **57267425** | **57277184** | **9** | **4.77** | **9.1E-07** |
|  | **GABRB3** | **2562** | **15** | **26788693** | **27018935** | **680** | **1.29** | **9.8E-02** |
|  | **CHRNA4** | **1137** | **20** | **61974665** | **61992695** | **50** | **2.93** | **1.7E-03** |
| closing | PODN | 127435 | 1 | 53527724 | 53551174 | 120 | 1.43 | 7.6E-02 |
|  | GCLM | 2730 | 1 | 94352590 | 94375012 | 63 | -0.59 | 7.2E-01 |
|  | BCAN | 63827 | 1 | 156611740 | 156629320 | 28 | -0.67 | 7.5E-01 |
|  | ADAMTS4 | 9507 | 1 | 161159538 | 161168845 | 20 | 0.40 | 3.4E-01 |
|  | TNR | 7143 | 1 | 175291935 | 175712752 | 1256 | 0.66 | 2.5E-01 |
|  | **RTN4** | **57142** | **2** | **55199325** | **55277734** | **227** | **3.76** | **8.4E-05** |
|  | **MME** | **4311** | **3** | **154797436** | **154901518** | **221** | **1.86** | **3.2E-02** |
|  | VCAN | 1462 | 5 | 82767493 | 82878122 | 277 | -0.28 | 6.1E-01 |
|  | HAPLN1 | 1404 | 5 | 82934017 | 83016896 | 228 | -0.31 | 6.2E-01 |
|  | **HLA-A** | **3105** | **6** | **29910247** | **29913661** | **5** | **2.24** | **1.3E-02** |
|  | HLA-C | 3107 | 6 | 31236529 | 31239855 | 3 | -0.49 | 6.9E-01 |
|  | **HLA-B** | **3106** | **6** | **31321649** | **31324989** | **99** | **6.66** | **1.4E-11** |
|  | GCLC | 2729 | 6 | 53362139 | 53409927 | 158 | -0.45 | 6.7E-01 |
|  | PILRB | 29990 | 7 | 99933688 | 99965454 | 59 | 0.91 | 1.8E-01 |
|  | CSGALNACT1 | 55790 | 8 | 19261672 | 19540261 | 1155 | 1.28 | 9.9E-02 |
|  | HAS2 | 3037 | 8 | 122625271 | 122653630 | 65 | 0.42 | 3.4E-01 |
|  | LYNX1 | 66004 | 8 | 143845756 | 143859640 | 42 | -1.01 | 8.4E-01 |
|  | MMP8 | 4317 | 11 | 102582526 | 102595685 | 72 | 0.73 | 2.3E-01 |
|  | MMP3 | 4314 | 11 | 102706528 | 102714342 | 22 | -0.92 | 8.2E-01 |
|  | PTS | 5805 | 11 | 112097088 | 112104696 | 16 | 0.64 | 2.6E-01 |
|  | **NCAM1** | **4684** | **11** | **112831969** | **113149158** | **933** | **4.28** | **9.5E-06** |
|  | ADAMTS8 | 11095 | 11 | 130274818 | 130298539 | 81 | -2.00 | 9.8E-01 |
|  | ADAMTS15 | 170689 | 11 | 130318869 | 130346539 | 65 | -1.20 | 8.9E-01 |
|  | **OTX2** | **5015** | **14** | **57267425** | **57277184** | **9** | **4.77** | **9.1E-07** |
|  | ACAN | 176 | 15 | 89346674 | 89418585 | 220 | -0.44 | 6.7E-01 |
|  | HAPLN3 | 145864 | 15 | 89420519 | 89438770 | 84 | 0.18 | 4.3E-01 |
|  | MMP15 | 4324 | 16 | 58059282 | 58080805 | 41 | 1.13 | 1.3E-01 |
|  | **HAS3** | **3038** | **16** | **69139467** | **69152622** | **23** | **1.66** | **4.9E-02** |
|  | DLG4 | 1742 | 17 | 7093209 | 7123369 | 50 | -1.46 | 9.3E-01 |
|  | MBP | 4155 | 18 | 74690789 | 74844774 | 733 | 1.12 | 1.3E-01 |
|  | **PTPRS** | **5802** | **19** | **5205519** | **5340814** | **489** | **4.58** | **2.4E-06** |
|  | **ICAM5** | **7087** | **19** | **10400655** | **10407454** | **23** | **2.62** | **4.4E-03** |
|  | **NCAN** | **1463** | **19** | **19322782** | **19363061** | **90** | **1.92** | **2.8E-02** |
|  | HAPLN4 | 404037 | 19 | 19366450 | 19373596 | 20 | 1.40 | 8.1E-02 |
|  | MAG | 4099 | 19 | 35782989 | 35804710 | 51 | 0.33 | 3.7E-01 |
|  | HAS1 | 3036 | 19 | 52216365 | 52227221 | 45 | 0.14 | 4.4E-01 |
|  | LILRB3 | 11025 | 19 | 54720147 | 54726959 | 26 | 0.28 | 3.9E-01 |
|  | LILRB1 | 10859 | 19 | 55128629 | 55149007 | 139 | 1.42 | 7.7E-02 |
|  | **MMP24** | **10893** | **20** | **33814539** | **33864804** | **91** | **3.00** | **1.3E-03** |
| expression | NGF | 4803 | 1 | 115828537 | 115880857 | 188 | -0.46 | 6.8E-01 |
|  | KCNK2 | 3776 | 1 | 215178885 | 215410436 | 728 | -0.33 | 6.3E-01 |
|  | **STAT1** | **6772** | **2** | **191833762** | **191878976** | **106** | **2.45** | **7.1E-03** |
|  | **CREB1** | **1385** | **2** | **208394616** | **208470284** | **125** | **2.99** | **1.4E-03** |
|  | TNF | 7124 | 6 | 31543350 | 31546112 | 4 | 4.27 | 9.8E-06 |
|  | **GRIN2A** | **2903** | **16** | **9847265** | **10276611** | **1908** | **2.90** | **1.8E-03** |
|  | DLG4 | 1742 | 17 | 7093209 | 7123369 | 50 | -1.46 | 9.3E-01 |
|  | PVALB | 5816 | 22 | 37196745 | 37215517 | 68 | -0.03 | 5.1E-01 |
| *Note.* Bolded rows indicate genes significant associated with depression risk at *p<0.05* in the gene-level results. | | | | | | | | |

Figure S1. A visual example of d-QTLs: SNP-level genotypes shaping the temporal patterns of gene expression in the medial prefrontal cortex.

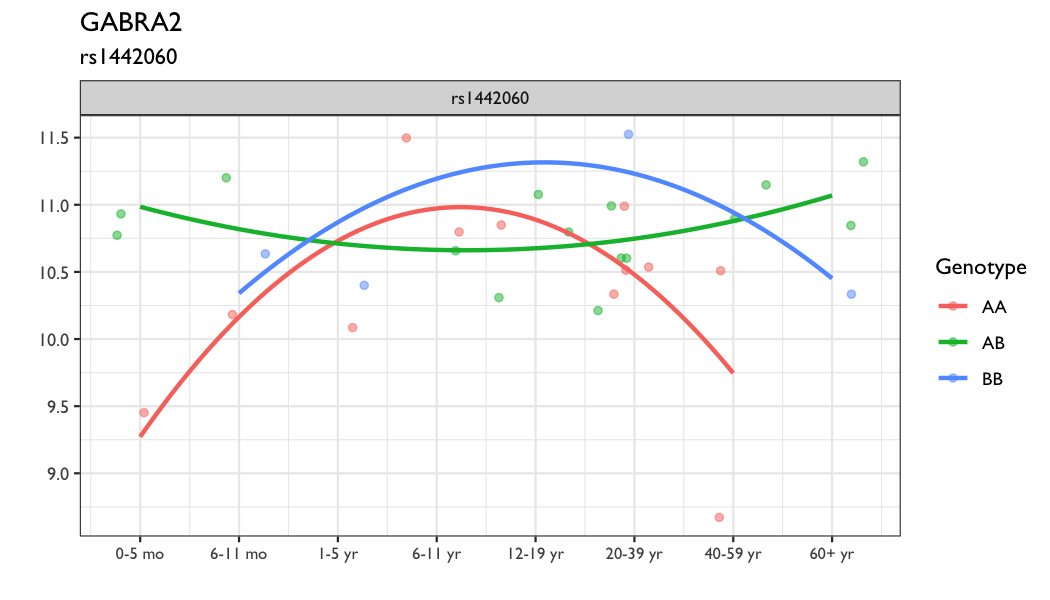

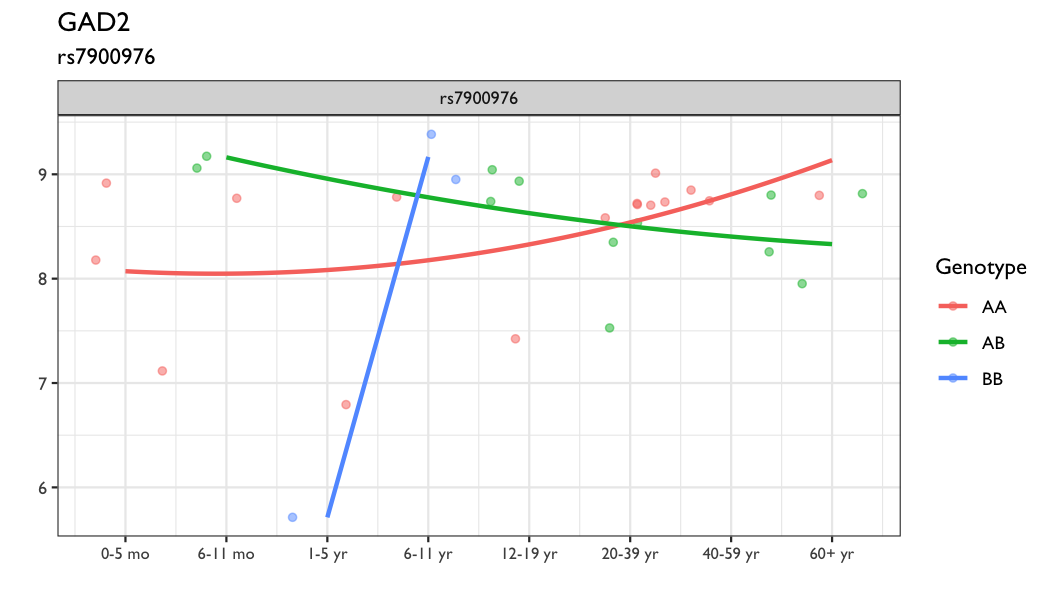

The developmental patterns of gene expression at *GABRA2* and *GAD2* were displayed above, stratifying by genotype at two SNPs that shaped gene expression trajectories. Genotype was modeled as a categorical variable and developmental stage was modeled with both a linear and a quadratic effect to account for the previously observed nonlinearity of timing effect while keeping the model parsimonious. An F-test was then performed to compare the model with the interaction terms included and the model without the interaction terms. All models adjusted for two principal components representing genetic ancestry.

Number of samples available varied across genotypes. For example, for GAD2, only two samples were available for minor allele homozygotes. We took these limitations into account when interpreting the trajectories across the life course.
